# Effects on hippocampal activity following 5-HT4 receptor agonism in unmedicated patients with depression: the RESTAND study

**DOI:** 10.1101/2025.08.29.25333049

**Authors:** Angharad N. de Cates, Amy L. Gillespie, Jessica Scaife, Marieke A.G. Martens, James Carson, Beata Godlewska, Wendy Howard, Anutra Guru, Philip J. Cowen, Catherine J. Harmer, Susannah E. Murphy

## Abstract

Cognitive impairment is a common but under-treated feature of Major Depressive Disorder (MDD). Preclinical and early human studies suggest that 5-HT_4_ receptor (5-HT_4_R) agonists rapidly improve learning and memory, consistent with this receptor’s role in hippocampal neuroplasticity. However, their effects in clinically depressed patients, remain unexplored.

In this double-blind, randomised study, 52 right-handed, unmedicated individuals with MDD received 6-9 days of the 5-HT_4_R agonist PF-04995274 (15mg, once daily) or placebo. Participants subsequently underwent fMRI scanning during a memory encoding task and completed behavioural measures of auditory verbal learning and spatial working memory.

Compared to placebo, PF-04995274 significantly increased activity in the hippocampus (ROI analysis) in response to novel versus familiar images, particularly in the left hemisphere. Whole brain analysis also revealed greater activation in the left inferior parietal lobule, a key region for memory processing. In contrast with previous studies using the 5-HT_4_R agonist prucalopride, PF-04995274 had limited effects on behavioural measures of memory.

The results demonstrate that short term 5-HT_4_R agonism enhances hippocampal and parietal activity during memory encoding in patients with depression. This replicates and extends previous findings in healthy volunteers using prucalopride, and is consistent with preclinical evidence establishing a key role for 5-HT_4_Rs in hippocampal-dependent learning and memory. This translational evidence supports a role for 5-HT_4_R activation in modulating memory-related brain circuits in MDD and highlights its therapeutic potential for treating cognitive symptoms of depression.

## Introduction

Depression is a leading cause of disability, affecting around 280 million people worldwide^1^. Conventional antidepressants, such as selective serotonin reuptake inhibitors (SSRIs) are not effective in treating all symptoms of depression. In particular, SSRIs do not adequately treat cognitive symptoms of depression and many patients have enduring problems with memory, concentration, attention and executive function following remission^2^. These symptoms occur in most clinical cases of depression (approximately 85-94%) and have been shown to remain in over 70% of patients with depression who have partially responded to SSRI treatment^3, 4^. Such residual symptoms are associated with greater risk of relapse, increased patient perception of disability, and a poorer prognosis in psychosocial and occupational outcomes^5, 6^. The development of treatments that target the cognitive symptoms of depression is therefore a key public health priority.

The serotonin 4 receptor (5-HT_4_R) is a promising novel target for the treatment of cognition. 5-HT_4_ Rs are widely expressed in regions related to mood and cognition in the human brain^7–9^, including the hippocampus, amygdala, nucleus accumbens, and medial prefrontal cortex. Preclinical work using animal models of depression and cognition has highlighted that 5-HT_4_R agonists act rapidly to improve performance on tasks of learning and memory^10–14^, including in animal models of depression^15^. These pro-cognitive effects are thought to be mediated via multiple mechanisms, such as increased hippocampal spine growth^16^, rapid potentiation of basal serotonin cell firing in the dorsal raphe nucleus^17^, stimulation of acetylcholine release^18^, modulation of glutamate transmission^19^, and the production of brain derived neurotrophic factor (BDNF)^20^.

Consistent with this preclinical work, the 5-HT_4_R agonist, prucalopride, has a pro-cognitive profile in healthy young adult volunteers^21–23^. A single dose of prucalopride is sufficient to improve performance across three separate measures of learning and memory, including declarative memory, reward learning and emotional memory^21^. A longer (six day) administration of prucalopride has also been shown to increase hippocampal activity during a memory task and improve accuracy of memory recall^22^. This translation of the pro-cognitive effects of 5-HT4R agonism from animal models to healthy human models represents an important step in establishing the potential of this receptor as a target for the treatment of cognition.

Using healthy participants for early mechanistic work in humans allows characterisation of neurocognitive effects of a novel agent without confounding due to symptom change. However, it is not yet known the extent to which these effects of 5-HT_4_R agonism generalise to a clinical population. Importantly, there have been no previous studies examining the neuropsychological effects of 5-HT_4_R agonists in individuals with psychiatric illness, a necessary step in further assessing the therapeutic potential of these agents.

To address this gap, the current study tested the effects of 5-HT_4_R agonism on learning, memory and hippocampal activity in a group of unmedicated patients with Major Depressive Disorder compared to placebo. The 5-HT_4_R agonist PF-04995274 was used; a highly selective 5-HT_4_R partial agonist initially developed as a potential therapeutic agent in Alzheimer’s Disease^24^, which was made available for the current study by Pfizer through the UK Medical Research Council Industry Asset Sharing Initiative. We hypothesised that, consistent with the effects of 5-HT_4_R agonism previously reported in healthy volunteers, PF-04995274 would improve memory performance and increase activation of the hippocampus during a memory task. Such a finding would represent a critical translation of the previous findings into a clinical population and further demonstrate the potential of the 5-HT_4_R as a target for the treatment of the cognitive symptoms of depression.

## Materials and methods

### Overview of design

This double-blind, randomised, between-group experimental medicine study assigned participants with un-medicated Major Depressive Disorder (DSM-5) to one of three groups: highly-selective 5-HT_4_R partial agonist PF-04995274 (15mg), or placebo, or the selective serotonin reuptake inhibitor citalopram (20mg), with stratification by gender. The study had two primary outcome measures: the effects of PF-04995274 on (i) memory processing and (ii) emotional processing respectively. It was powered to examine differences between the PF-04995274 and placebo group as the primary comparison, with the citalopram group included in the design as a secondary active comparator to placebo for emotional cognition analyses (due to the well-established effect of SSRIs on emotional processing). The outcomes related to memory (the fMRI memory task and associated secondary behavioural measures of learning and memory) are reported here and emotional processing outcomes are reported elsewhere^25^. As the focus of this report is to characterise the cognitive effects of 5-HT_4_R agonism (PF-04995274) versus placebo, we therefore report this comparison as principal analyses (see Supplementary Material for main outcomes comparing citalopram and placebo groups for reference).

### Participants

Right-handed participants between 18 and 61 years old were recruited between August 2018 and July 2022. Participants were of either sex, fluent in English, and were screened for contraindications to serotonergic medication. All participants met DSM-5 criteria for Major Depressive Disorder at screening, as determined by a psychiatrist using the Structured Clinical interview for DSM-5 (SCID). Other criteria for inclusion included: being willing and able to give consent; not receiving antidepressant drug treatment or face-to-face psychological treatment for the last six weeks; and, where relevant, using a highly effective method of contraception from study enrolment until 30 days after study treatment. Participants who had failed to respond to an antidepressant or who had been given electroconvulsive therapy (ECT) in the current depression episode were excluded, as well as those with a diagnosis of bipolar disorder or at high risk of suicide (see Table S1 for a complete list of all exclusion criteria). The study was approved by the South Central Research Ethics Committee (18/SC/0076) and pre-registered with clinicaltrials.gov (NCT03516604). Participants provided written informed consent and received £200 compensation for their time.

### Intervention and randomisation

PF-04995274 and identically matched placebo tablets were provided by Pfizer. Citalopram and placebo were sourced and encapsulated by Cardiff and Vale University Health Board, St Mary’s Pharmaceutical Unit to also ensure identical matching of the placebo and citalopram. Participants were asked to take four pills daily in the morning. The PF-04995274 group received three tablets of PF-049952 (15mg in total) plus one placebo capsule, the citalopram group received one citalopram capsule (20mg) plus three placebo tablets, and the placebo group received three placebo tablets and one placebo capsule. The Oxford Health NHS Foundation Trust Pharmacy held the Randomisation List and were responsible for storing and dispensing the study medication. Participants were required to take the medication for 7 days, plus up to an additional 2 days, where necessary, to facilitate study visit and scan bookings. The PF-04995274 dosing (15 mg for ≥6 days) was previously established as safe^26^, with a half-life (t_1/2_) of around 30 hour and >80% brain 5-HT_4_ R occupancy after first dose in healthy volunteers^24^.

### Study visits and procedure

The study involved four visits to the Warneford Hospital, University of Oxford Department of Psychiatry: (i) Screening (optionally online), (ii) First Dose, (iii) Research Visit One (fMRI scan, days 6-9), and (iv) Research Visit Two (behavioural tasks and HAM-D, days 7-9). Screening included medical and psychiatric evaluation, vital signs assessment, ECG, blood tests, drug screening, and pregnancy testing where applicable. The first dose was administered under medical supervision, with vital signs monitored for three hours before participants were allowed to leave. Participants took remaining doses at home up until and including the date of Research Visit Two, receiving phone checks on days two and four and daily medication reminders. During Research Visit One, participants completed a 3T MRI scan at the Oxford Centre for Human Brain Activity: an fMRI memory encoding task and arterial spin labelling (ASL) (reported here), an fMRI emotional faces task (reported elsewhere^25^) and a resting state scan. During Research Visit Two, participants completed a battery of tasks, including the Auditory Verbal Learning Task and the Oxford Memory Task to assess memory (reported here).

### Sample Size

The target sample size was 25 participants per group. This powered the study adequately to detect an effect size of the same magnitude as that reported in our previous study^22^ showing an effect of 5- HT_4_R agonism on hippocampal activity (using the same hippocampal region of interest approach as used here) with 90% power (G^*^Power, previous effect size (f) = 0.37, with an α=0.05, to give 90% power (1-β)).

### Questionnaire measures

Participants completed the following self-report questionnaires at Screening to obtain measures of trait anhedonia, anxiety, and personality: Snaith–Hamilton Pleasure Scale (SHAPS)^27^, Spielberger State-Trait Anxiety Inventory, Trait Version (STAI-T)^28^, and Eysenck Personality Questionnaire^29^. Affect and state anxiety were also measured pre- and post-imaging (Research Visit 1): affect using the Positive and Negative Affect Scale (PANAS)^30^ and the visual analogue scale (VAS)^31^; anxiety using the Spielberger State-Trait Anxiety Inventory, State Version (STAI-S)^28^. Side effects were measured at Screening and pre- and post-imaging using a scale where participants rated their experience for potential side effects^32^. Participants were also assessed on an observer-rated Hamilton Rating Scale for Depression (HAM-D) by trained researchers and completed a self-report measure of depressive symptoms (Beck Depression Inventory-II (BDI))^33^ at Screening and Research Visit Two. At the end of Research Visit Two, participants guessed their drug allocation with a forced-choice question.

### Memory encoding fMRI task

The memory encoding fMRI task, programmed in Presentation (Neurobehavioral Systems; https://www.neurobs.com) (see Figure 1(A)), was designed to stimulate implicit visual memory recognition in the scanner for images that participants had seen before (familiar images), compared to images that were new to participants in the scanner (novel images) where implicit encoding should occur. See Figure 1(A), Supplementary Material and ^22^ for further task details.

**Figure 1.**
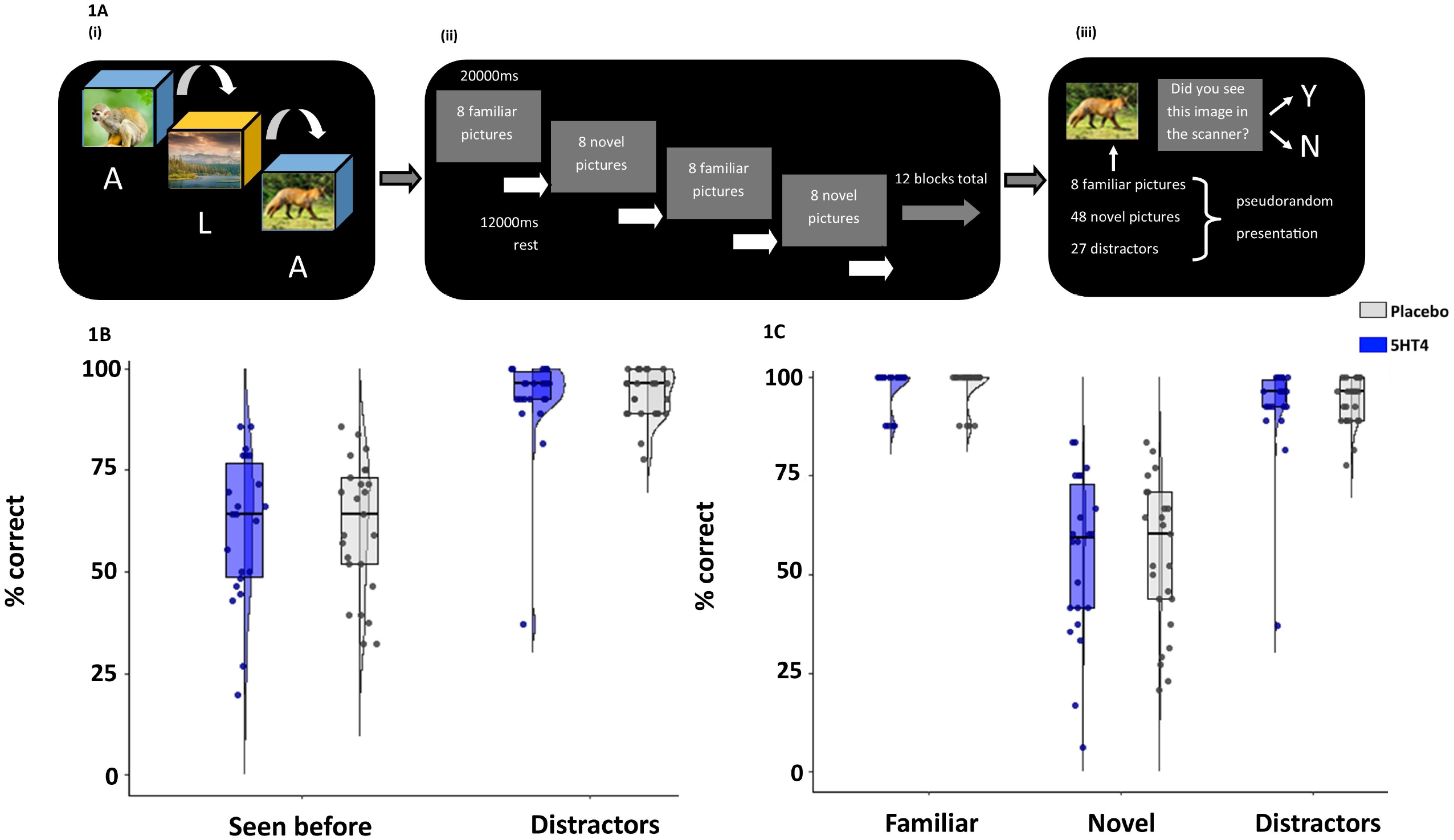
Post-scan recall task (A) results (percentage total correct at identifying image type) divided by group (placebo versus PF-04995274 (5-HT_4_)): (B) seen before (novel+familiar) vs. distractors; (C) novel vs. familiar vs. distractors. (A) As previously detailed (de Cates et al., 2021): (i) Prior to the scan, eight “familiar” pictures were shown to participants chosen (four animals and four landscapes). These images were shown eight times each in pseudorandom order on a computer screen, with each image displayed for 3250 ms, a 500 ms interval between images, and a 5000 ms interval (featuring a fixation cross) between blocks. While viewing the images, participants used a 2-button response to categorise each as either animal or non-animal (A or L), and their understanding of the task was confirmed. (ii) During the scan, blocks of both familiar and novel images were presented: 6 blocks of familiar images and 6 blocks of novel images. Each block lasted 20,000 ms, with individual images shown for 2000 ms and a 500 ms interval between images. A rest period of 12000 ms with a fixation cross followed each block (making a total of 12 rest blocks). Participants continued to indicate via a 2-button response whether each image was an animal or non-animal. Compliance and accuracy was monitored by scanner operators. Responses were automatically recorded in a text file by the software. (iii) After scanning, participants completed a recall test in which they identified which images they had seen during the scanning task and which were distractor images. (B) Grey = placebo, Blue = 5-HT_4_ for **(B)** seen before (novel + familiar) vs. distractors; **(C)** novel vs. familiar vs. distractors. Box plots represent the interquartile range (IQR) with central line depicting the median. Whiskers represent +/− 1.5 IQR. Data points and half violin plots depict the data distribution.

### Auditory verbal learning task (AVLT)

Participants learned a list of 15 concrete nouns (List A) across five immediate recall trials, then were asked to recall immediately a second unrelated list (List B). They were later asked to recall List A both after 2 minutes, and then again following a 20-minute delay, with accuracy, repetitions, and intrusions recorded (see Figure 4(A)). Finally, they completed a recognition task involving List A words and distractors, with hits and false alarms measured.

### Oxford Memory Task (OMT)

The OMT^34^ is a spatial working memory and item-location binding task designed for clinical populations. It contained 2 practice blocks with 10 trials for each and followed by 3 test blocks with 40 trials for each. In each trial, either 1 or 3 fractal objects were randomly located on the screen for 1 or 3s, respectively. Participants were asked to remember both the fractals and the locations. A blank delay screen was then displayed, followed by a test array with two vertically presented fractals, in which one was the target and the other was a foil. Participants were required to pick the target fractal and drag it to the remembered location. OMT performance was measured by accuracy (the proportions of trials in which the targets were correctly chosen), identification time (reaction time during which participants picked the object from the test array), and localisation time (reaction time during which participants dragged and placed the object).

## Demographic and behavioural data analysis

Behavioural data and questionnaires were pre-processed and analysed using SPSS (version 29, IBM) and R (version 4.3.3). Graphs were produced using GraphPad Prism (version 9) and R (version 4.3.3). A repeated-measures analysis of variance (ANOVA) was used to analyse group differences in self-report measures and behavioural performance in the AVLT, OMT, and fMRI experimental task. Levene’s test (t-tests) and the Greenhouse–Geisser procedure (ANOVAs) were used where appropriate. A p-value of less than 0.05 was used to denote statistical significance. Partial eta squared is reported as a measure of effect size. Sex, age and HAM-D scores using ANCOVAs were considered in sensitivity analyses as appropriate.

### MRI data acquisition and analysis

Blood-oxygenation-level-dependent (BOLD) fMRI and T1-weighted anatomical images were acquired using a 3-Tesla Siemens Prisma scanner, equipped with a 32-channel head matrix coil (Siemens, Erlangen, Germany). Foam padding and a head restraint were used to control head movement. Further details of fMRI and structural MRI acquisition can be found in the Supplementary Material. The full acquisition protocol, along with radiography procedure, is available from the Open WIN MR Protocols database here: 10.5281/zenodo.6107724.

Imaging data were analysed with FSL (www.fmrib.ox.ac.uk/fsl). fMRI data were pre-processed and analysed using FEAT (FMRI Expert Analysis Tool), version 6.0.4, part of FSL (FMRIB’s SoftwareLibrary; www.fmrib.ox.ac.uk/fsl). Full details regarding pre-processing are in the Supplementary Material. In the first-level analysis, individual activation maps were computed using the general linear model with local autocorrelation correction. Two explanatory variables were modelled: “novel” and “familiar” images. Temporal derivatives were included in the model. Variables were modelled by convolving each block with a haemodynamic response function with a standard deviation of 3s and mean lag of 6s. At the whole-brain level, the following model was constructed: (1) novel vs. baseline; (2) familiar vs. baseline; (3) novel > familiar; (4) novel < familiar. In the second-level analysis, whole-brain individual data were combined at a group level (placebo vs. 5-HT_4_) using a mixed-effect analysis. Cerebral blood flow and grey matter maps were included as pre-specified covariates of no interest. Details of the acquisition and analyses for these are in Supplementary Material. We also considered other covariates (sex, age, HAM-D score) similar to behavioural analyses. Groups were compared in the following manner: (1) placebo > 5-HT_4_, (2) 5-HT_4_ > placebo, (3) placebo mean, (4) 5-HT_4_ mean, and (5) mean of all participants. Brain activations showing significant group differences were identified at the whole-brain level using cluster-based thresholding (Z > 3.1, familywise error (FWE) *p*<0.05 corrected). Significant interactions from whole-brain analyses were further explored by extracting percentage BOLD signal change for each type of contrast. As the hippocampus was a particular focus, it was pre-specified as a region of interest (ROI). A functional ROI mask was created for the left and right hippocampus by multiplying mean activation for each contrast of interest (on already corrected whole-brain data as above) for all participants by the structural Harvard-Oxford subcortical atlas mask at 50% threshold. Percentage BOLD signal change was subsequently extracted for each contrast in each hemisphere.

## Results

### Participants

In total, 55 participants were recruited to the placebo and PF-04995274 groups and completed fMRI data collection (26 placebo: 27 PF-04995274) between 1^st^ August 2018 and 19^th^ July 2022. Three participants withdrew earlier in the study: one due to abdominal pain and one due to migraine (both placebo); one due to multiple symptoms including sleep disturbance, headache and brain fog (day 4 – PF-04995274). All symptoms resolved with 24 hours of discontinuing their allocated intervention. One participant (PF-04995274) was excluded from fMRI analyses prior to unblinding for significant movement. For the remaining 52 participants (26:26; placebo: PF-04995274), no individual participant had absolute movement greater than one voxel or relative movement more than ½ voxel, and there were no significant differences between groups in terms of movement (absolute: *p*=0.07; relative: *p*=0.71). Mean medication length in days was similar for both groups (PF-04995274 mean (SD) = 8.00 (0.94); Placebo mean (SD) = 7.96 (0.72).

Participants who had MRI contra-indications or exclusions but completed the secondary behavioural measures (AVLT and OMT) were included for these tasks (28 placebo: 31 PF-04995274). For full details including a CONSORT diagram, see Figure S1 and ^25^.

At baseline, the fMRI subset groups were well matched in terms of demographics (e.g. age, sex, BMI, first language, years of education, handedness) and clinical presentation (e.g. substance use, clinician-rating depression (HAM-D), self-assessed mood (BDI), anhedonia, anxiety); see Table 1.

**Table 1.** Baseline demographics and questionnaire results for RESTAND study (placebo versus PF-04995274 (5HT_4_))

| Variable (measure) | Measurement |  | Placebo N=26 | PF-04995274 N=26 | P value | Odds ratio |
| --- | --- | --- | --- | --- | --- | --- |
| **Sex** | *Number* | *Male* | 11 | 10 |  | Ref |
|  |  | *Female* | 15 | 16 | 1.00 | 1.17<sup>1</sup><br>(0.39-3.56) |
| **Age** | *Mean in years (SD; range)* |  | 31.8 (10.1; 18.9-55.4) | 37.0 (13.2; 18.9-60.7) | 0.11 |  |
| **Years of education\*** | *Mean (SD)* |  | 17.2 (2.71) | 16.7 (2.50) | 0.64 |  |
| **First language** | *Number* | *English* | 22 | 16 |  | Ref |
|  |  | *Not English* | 4 | 10 | 0.12 | 3.44 (0.91-13.0)<sup>2</sup> |
| **Handedness (EHI)** | *Mean % right (SD)* |  | 81.6 (2.66) | 81.1 (1.86) | 0.43 |  |
| **Smoking\*<sup>3</sup>** | *Mean cigarettes / day (SD)* |  | 0.06 (0.30) | 0.04 (0.20) | 0.76 |  |
| **Caffeine\*** | *Mean cups / day (SD)* |  | 2.80 (1.72) | 2.40 (1.64) | 0.40 |  |
| **Body mass\* index** | *Mean (SD)* |  | 25.4 (4.21) | 24.9 (4.64) | 0.69 |  |
| **BDI** | *Mean (SD)* |  | 25.7 (10.06) | 23.7 (10.66) | 0.49 |  |
| **HAM-D** | *Mean (SD; range)* |  | 16.9 (3.02; 13-25) | 15.9 (2.32; 12-21) | 0.17 |  |
| **Anhedonia (SHAPS)** | *Mean (SD)* |  | 4.08 (3.12) | 4.19 (2.68) | 0.89 |  |
| **Anxiety (STAI-T)** | *Mean (SD)* |  | 59.8 (7.22) | 58.8 (9.10) | 0.68 |  |
| **Personality (EPQ)** | *Mean (SD)* | *Neuroticism/Stability* | 18.0 (4.06) | 17.4 (4.55) | 0.65 |  |
|  |  | *Psychoticism/Socialisation* | 3.38 (2.68) | 4.62 (2.90) | 0.12 |  |
|  |  | *Extroversion/Introversion* | 9.12 (5.56) | 8.31 (4.92) | 0.58 |  |
|  |  | *Lie/Social desirability* | 8.23 (5.05) | 7.46 (3.71) | 0.53 |  |

<sup>1</sup> OR of being a female in the 5-HT<sub>4</sub> group, compared to males
<sup>2</sup> OR of being non-native English speaker in the PF-04995274 group, compared to English speaker
<sup>3</sup> \* = missing data (1 missing from placebo group for substances; 1 missing from placebo and 2 from 5-HT<sub>4</sub> groups for BMI; 7 missing from each of placebo and PF-04995274 groups for education)

Mean HAM-D at baseline indicated that most participants were in the mild to moderate depression range. Both groups contained a similar number of participants with ongoing physical health medication, and no participant reported serious medical illness (see Tables S2 and S3). For tables including participants with secondary behavioural measures see ^25^.

### Responses for side effects and randomisation guesses

At the end of the study, participants were asked to guess their study randomisation. Participants in the PF-04995274 group were significantly less likely to correctly identify their randomisation compared to placebo (odds ratio (Pearson chi-square), *p*=0.043). Of those who guessed incorrectly in the PF-04995274 group (9/26), two thirds suggested placebo and one third suggested citalopram (see Table S4). A further six placebo and 12 5-HT_4_ participants reported they had “no idea” of their allocation.

### Behavioural results of the fMRI memory encoding task

All groups completed the pre-scan training in full and performed well on the in-scan accuracy task (t(50) = −0.35, *p*=0.73; placebo (M=98.5%, SEM=0.51), PF-04995274 (M=98.7%, SEM=0.26)), indicating that participants were engaging with the task as required.

In the post-scan recall test, there was no difference between groups in accuracy to identify images seen before (novel + familiar) versus distractors [*F*(1,45)=0.081, *p*=0.777, ηρ2<0.01; overall mean for placebo (M=77.2% SEM=1.82), PF-04995274 (M=76.5%, SEM 1.94)], or when we analysed that according to each category of image (novel/familiar/distractor) [F(1,45)=0.079, *p*=0.779, ηρ2<0.01, see Figure 1(B) and (C)]. There was no interaction between image type (novel/familiar/distractor) and group [F(1.36, 61.0)=0.019, *p*=0.94, ηρ2<0.01 (data missing for five participants (1 from placebo, 4 from PF-04995274 = 47 (25:22) participants included)]. Results were unchanged when age, HAM-D score and sex were included as covariates (see Table S6).

### Analyses assessing for potential confounds in fMRI analyses

Grey matter and cerebral perfusion were included as covariates in analyses as pre-specified. Group-level effects on grey matter and cerebral perfusion are detailed in Supplementary Material (Figure S2 and Table S7).

Activity between groups within the region of interest hippocampal mask did not vary as a function of HAM-D scores at baseline [F(1,49)=0.04, *p*=0.85, ηρ2=0.00], or age [F(1,49)=3.05, *p*=0.087, ηρ2=0.06]. However, activity within the mask did vary as a function of sex [F(1,49)=11.93, *p*<0.01, ηρ2=0.20]. Sex was therefore included as a covariate in region of interest analyses, as well as grey matter and perfusion as pre-specified. Results from whole-brain analyses were very similar when sex was incorporated into analyses in addition to grey matter and perfusion (see Supplementary Material).

### fMRI memory encoding task

## Main effect of task

Consistent with previous reports^35, 36^, the novel versus familiar contrast revealed increased BOLD fMRI signal intensity bilaterally in the hippocampus, parahippocampal gyrus, and temporal fusiform cortex. Similar to a previous study using prucalopride^22^, there was also increased activity to novel versus familiar pictures in association areas including the thalamus, regions in the frontal (inferior frontal gyrus, frontal pole, precentral gyrus, cingulate / paracingulate gyrus) and occipital lobes (lateral occipital cortex), cerebellum, basal ganglia (caudate, putamen) and amygdala (see Figure S3).

## Effect of treatment – region of interest analysis

In the hippocampal region of interest mask, there was a significant condition^*^hemisphere^*^group interaction [F(1,49)=4.41, *p*=0.04, ηρ2=0.08]. Post-hoc pairwise comparisons identified that this significant interaction was predominately driven by increased activity in the PF-04995274 group to novel compared to familiar images, particularly in the left hemisphere [left: estimated marginal mean (EMM) difference=0.08 (0.01-0.15), *p*=0.035; right: EMM difference=0.06 (0.00-0.11), *p*=0.050], which was less evident in the placebo group [left: EMM difference=0.01 (−0.06-0.08), *p*=0.789; right: EMM difference=0.06 (0.00-0.11), *p*=0.053].

There was no significant condition^*^group interaction for either hemisphere individually: [LHC novel vs. familiar: F(1,49)=1.80, *p*=0.19, np2=0.04; RHC novel vs. familiar: F(1,49)=0.00, *p*=0.99, np2=0.00]. Furthermore, there was no significant difference in activity between the two groups in response to both novel and familiar images (versus baseline) in the left and right hemisphere [F(1,49)=1.39, *p*=0.24, ηρ2=0.03; see Figure 2].

**Figure 2.**
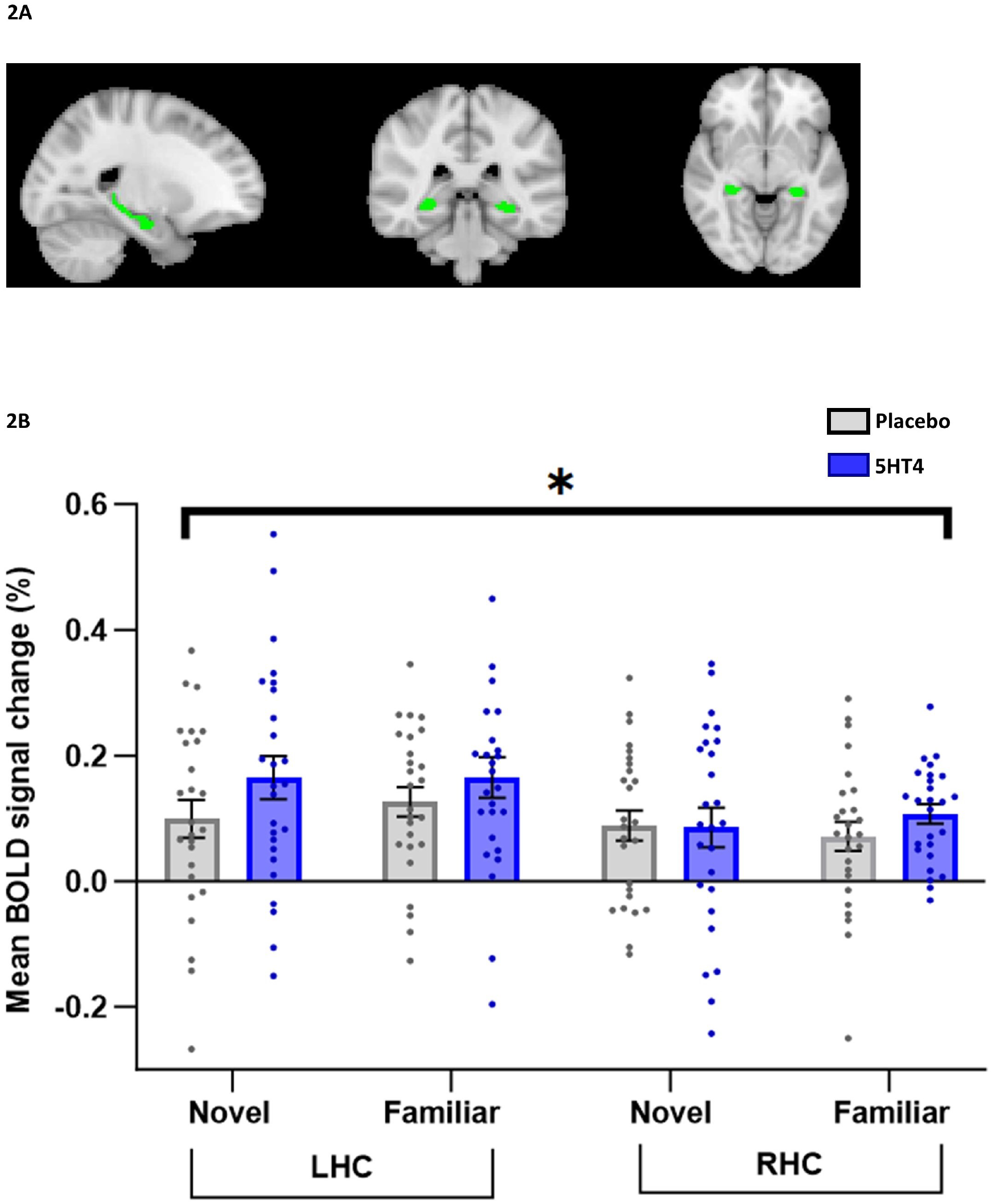
BOLD % signal change extracted from the functional left (LHC) and right (RHC) hippocampal mask in response to novel and familiar images by group (placebo vs. PF-04995274 (5-HT_4_)) (A) A functional ROI mask was created for the left and right hippocampus for each contrast of interest by multiplying mean activation for all participants by the anatomical mask at a 50% threshold. LHC and RHC mask in sagittal, coronal, and axial views created for novel and for familiar mean (novel shown at MNI −22, −36, −6 (green)) (B) Group mean of the BOLD percentage signal change extracted from the left and right hippocampal masks for novel and familiar images. Grey = placebo, Blue = 5-HT_4._ Error bars show standard error of the mean. ^*^ denotes significance within the ANOVA (p<0.05)

## Effect of treatment – whole brain analysis

There was increased activity in the PF-04995274 group compared with the placebo group in response to novel images versus baseline in the left inferior parietal lobule [5-HT_4_ > placebo, Z=4.19, p<0.007, peak voxel location: X=−36, Y=−88, Z=32, cluster size = 151 voxels (Figure 3(A))]. Figure 3(B) represents the group-level extracted BOLD signal change for this activation cluster. There were no significant differences between the groups at a whole brain level for the novel > familiar contrast, novel < familiar contrast, or the familiar > baseline contrast. Whole-brain analyses were very similar when sex, baseline HAM-D score or age were included as regressors in the analyses (see Supplementary Material).

**Figure 3.**
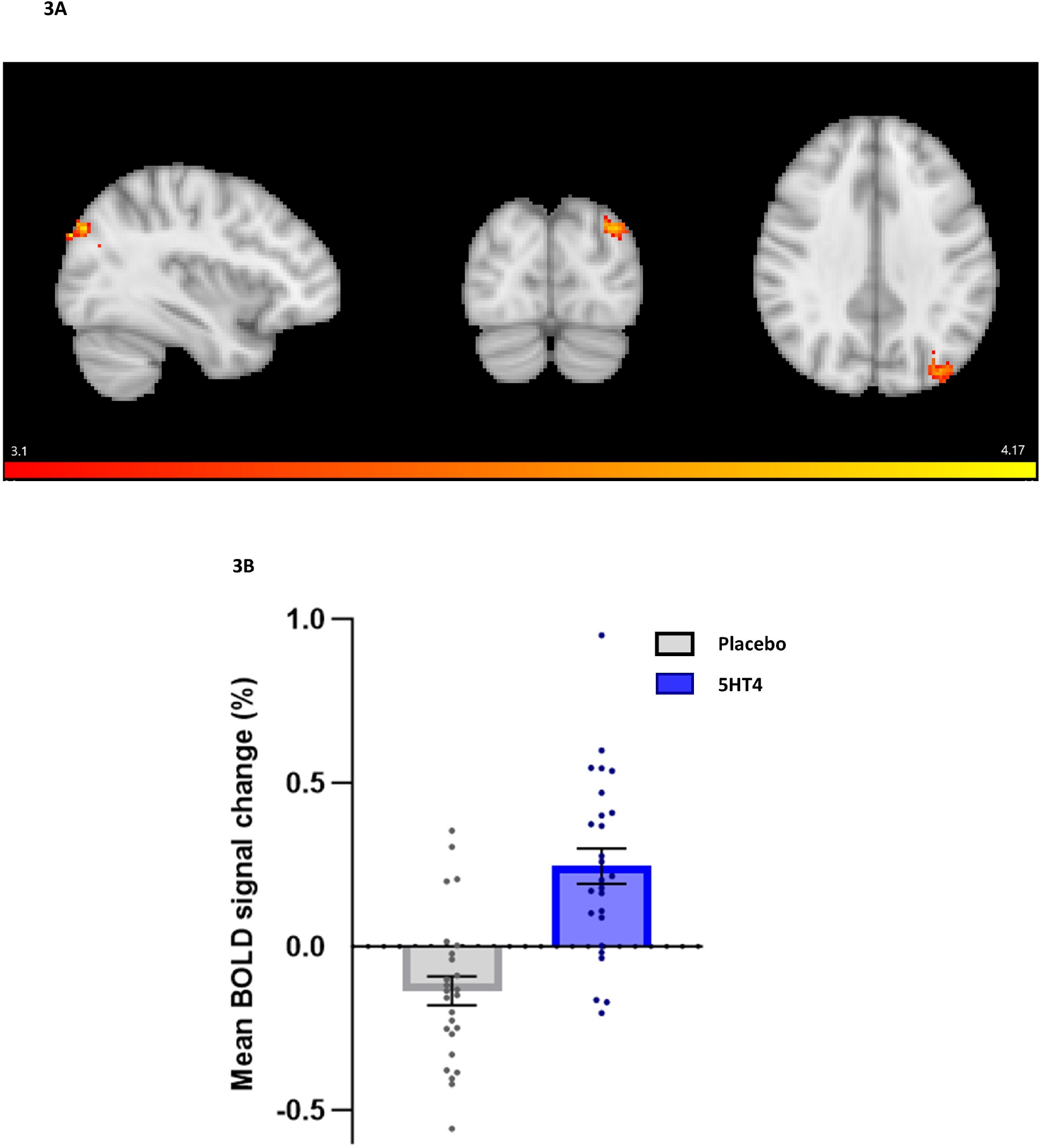
Whole-brain activation in response to novel images (compared to baseline) in PF-04995274 (5-HT_4_) versus placebo group. (A) Sagittal, coronal and axial images depicting significantly increased activation in the 5-HT_4_ group for the novel > baseline contrast in a left inferior parietal lobule cluster (peak voxel X=−36, Y=−88, Z=32, cluster size = 151 voxels). Images shown at MNI −36, −82, 34, thresholded at z > 3.1, p<0.05 corrected. Red to yellow colours identify increased in brain activation (B) Group mean of the BOLD percentage signal change extracted from the left lateral occipital cortex cluster (novel > baseline). Grey = placebo, Blue = 5-HT_4._ Error bars show standard error of the mean.

### Auditory Verbal Learning Task

Two PF-04995274 participants were missing AVLT data (placebo: PF-04995274 = 29:29) and data from one PF-04995274 participant was excluded prior to unblinding specifically for the long delay analysis due to the onset of menstrual cramps, which interrupted performance at the end of the task. There was a significant effect of block on word recall [F(7,392) = 202, p < 0.001, np2=0.78], indicating that in both groups participants’ recall of words from List A improved across the five acquisition blocks (Figure 4(B) and Table S8). However, there was no significant interaction between the block and group [F(7,392) = 0.35, p = 0.93, np2<0.01] or main effect of group [F(1,56) = 0.02, p = 0.89, np2<0.01]. There was also no group difference in the number of words accurately recalled after a short delay [F(1,56) = 0.00, p = 0.96] or a long delay [F(1,55) = 0.13, p = 0.72]. There were no differences between groups in terms of intrusions or repetitions, or recall of a separate set of words (List B) (all ps > 0.3, see Table S9).

**Figure 4.**
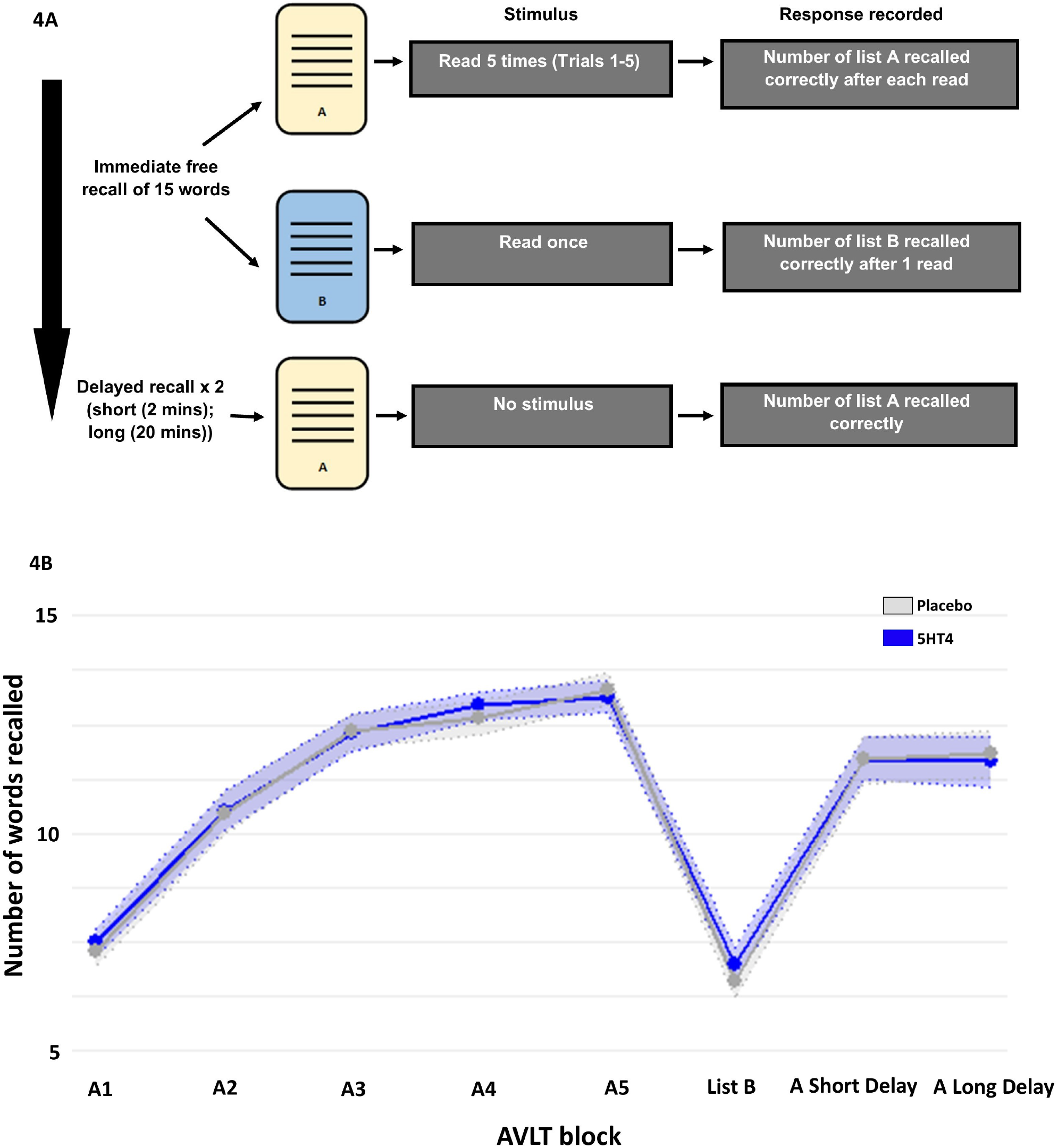
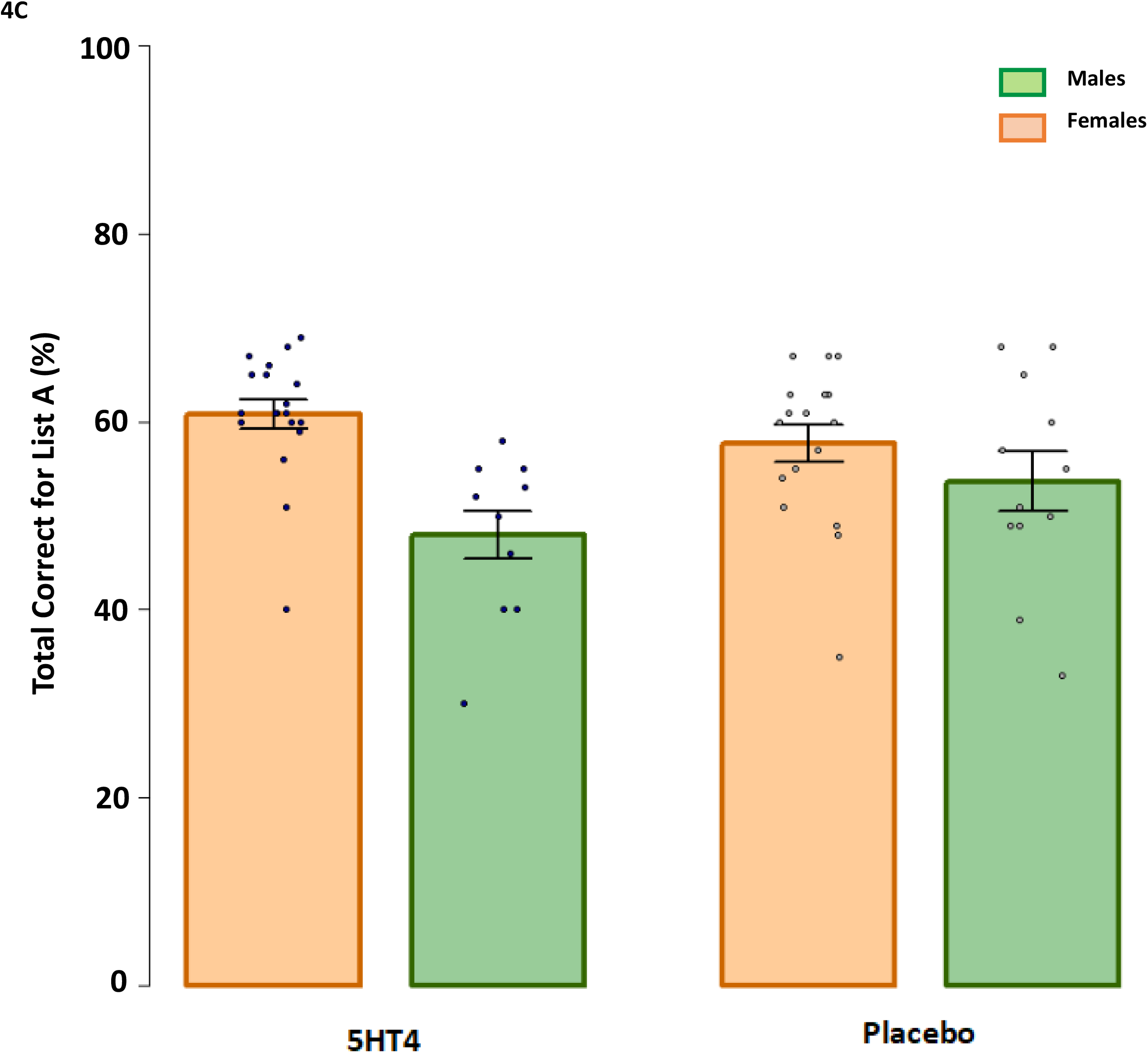
Scores on the Auditory Verbal Learning Task by group (placebo versus PF-04995274 (5-HT_4_)) (A) Participants were read a list of 15 concrete nouns (List A) and asked to immediately verbally recall as many items as they could. This was repeated five times before a list of unrelated words was presented (List B) and participants were again asked to recall them. Participants were then asked to recall List A immediately (short-delay) and after a delay of ~20 min (long-delay). Number of words correct, repetitions (correct words recalled more than once in the same acquisition trial) and intrusions (incorrect words not present in the list) were measured. Participants then completed a recognition task where they were required to indicate which of a list of words (15 List A words, 35 distractors) had previously been presented. Number of hits and false alarms were measured. (B) Word recall for each block shown individually as group mean (A1 = List A Trial 1; A2 = List A Trial 2; A Short Delay = List A Short Delay; A Long Delay = List A Long Delay). Shaded area = standard error of the mean. (C) Scores for List A total score subdivided by sex (Orange = Females; Green = Males). Error bars show standard error of the mean. Data points depict the data distribution.

There was greater total recall for females compared to males in the PF-04995274 group [mean (SD): Placebo females 57.6 (8.8), males 55.0 (10.3); PF-04995274 females 60.4 (6.7), males 46.7 (9.6), see Figure 4(C)]. This was associated with a significant interaction between sex and group [F(1,54) = 5.74, p = 0.02, np2=0.07]. See Table S10 for the effect of sex for other parts of the AVLT.

### Oxford Memory Task

One placebo participant was missing data for the OMT task (placebo: PF-04995274 = 28:31). There was no difference between groups in terms of accuracy [main effect of group (F(1,52.0)=1.06, *p* = 0.31, np2 = 0.02); fractals^*^group (F(1,58.4)=1.58, *p* = 0.21, np2 = 0.03)], or identification time [main effect of group (F(1,50.0)=0.09, *p* = 0.77, np2 < 0.01); fractals^*^group (F(1,57.7)=0.07, *p* = 0.78, np2 < 0.01)]. There was a borderline effect on localisation time [main effect of group (F(1,53.5)=2.57, *p* = 0.11, np2 = 0.05); fractals^*^group (F(1,59.7)=0.84, *p* = 0.36, np2 = 0.01)]. This main effect was driven by the 5-HT_4_ group having faster localisation time than the placebo group (see Table S12). When age and HAM-D score at study end were added as covariates, this main effect of group showed a significant difference between groups (age: F(1,52.4)=4.10, *p* = 0.04, np2 = 0.07; HAM-D: F(1,51.0)=3.71, *p* = 0.05, np2 = 0.07), although interactions between age / HAM-D score and group were not significant (ps > 0.05; see Figure S4).

### Subjective affect and anxiety and the effect of PF-04995274 on mood during the study

There were no significant differences in state anxiety or affect between the PF-04995274 and placebo group during the scan visit (all *p*s > 0.4; see Table S5).

As otherwise reported^25^, the PF-04995274 group had a significantly lower HAM-D scores than those participants receiving placebo at study end. This was also true in the subset of participants who completed the fMRI memory encoding task reported here [t(49) = 2.87, *p*=0.006; placebo follow-up HAM-D (M=15.4, SD=4.08), PF-04995274 follow-up HAM-D (M=12.2, SD=3.72); data missing for one PF-04995274 participant] (see Figure 5).

**Figure 5.**
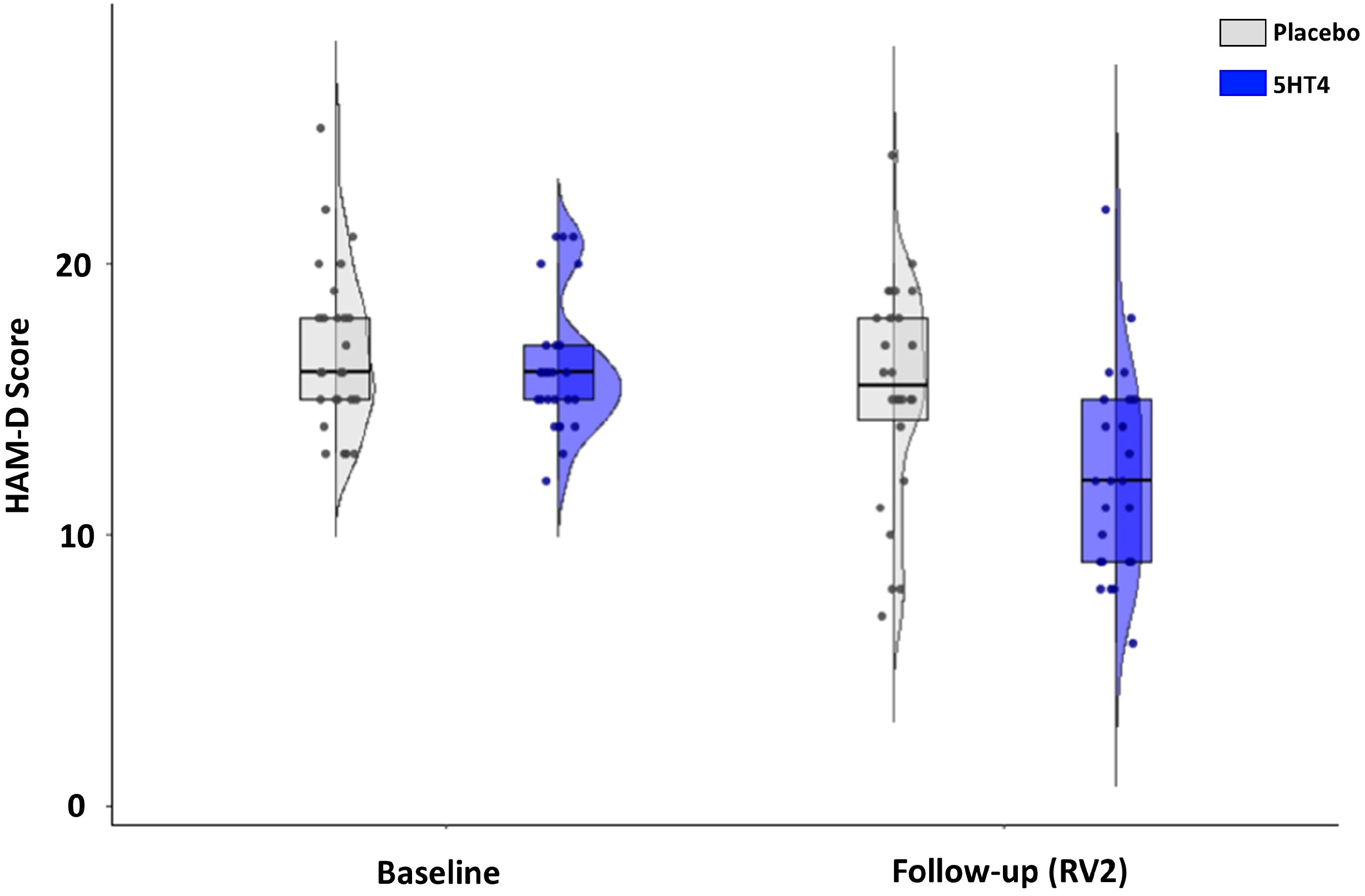
HAM-D change during study for fMRI subset of participants. Box plots represent the interquartile range (IQR) with central line depicting the median. Whiskers represent +/− 1.5 IQR. Data points and half violin plots depict the data distribution.

## Discussion

In unmedicated depressed volunteers, the novel 5-HT_4_R agonist, PF-04995274, increased neural activation in the context of a memory encoding task in two regions relative to placebo: (i) the hippocampus and (ii) the left inferior parietal lobule (IPL). This is consistent with previous work in animal models and healthy volunteers reporting increased hippocampal activity following 5-HT_4_R agonist administration. On behavioural measures of memory, PF-04995274 did not have a clear pro-cognitive effect, apart from a slight speeding of response on the Oxford Memory Task. This suggests it may have a more limited profile of effect than that previously reported with the 5-HT_4_R agonist, prucalopride.

In pre-specified region of interest analyses, this study replicated the finding that 5-HT_4_R agonism in humans increases hippocampal activity in the context of a memory task. Importantly, this study extended this finding from healthy volunteers^22^ into a clinical sample of depressed patients. The hippocampus is well-established as a memory encoding region, especially when rich mental imagery is involved^37^, and is closely connected to memory association areas, including the angular gyrus (part of the IPL)^38, 39^. 5-HT_4_ Rs are known to be expressed in the hippocampus^26^, and animal data has consistently demonstrated that agonism of these receptors plays an important role in facilitating hippocampal-dependent learning and memory. The hippocampus is a particularly important region within the context of depression^40, 41^; reduced hippocampal volume is a consistent and well-documented finding^42^, and is associated with markers of disease progression, including number of episodes, illness duration, and treatment resistance^43^. Depression is also associated with altered hippocampal function; for example, the hippocampus has been reported to have impaired functional connectivity with both the prefrontal cortex^44, 45^ and amygdala^46, 47^ in people with untreated depression, and these alterations in connectivity have been associated with depression severity^47^. The facilitation of hippocampal function by PF-04995274 in people with depression, seen in the current study, further lends weight to the idea that the 5-HT_4_R holds significant promise as a target for the treatment of depression.

In whole-brain analyses, participants receiving PF-04995274 had increased activity in the IPL for the novel > baseline contrast, which includes both the supramarginal gyrus and the angular gyrus. These whole-brain fMRI findings are similar to our healthy volunteer study^22^, in which we reported increased activity in the (right) angular gyrus following 6 days administration of the 5-HT4R agonist prucalopride (although here in response to familiar images). The angular gyrus is known to play a role in memory retrieval, and in particular in distinguishing objects that were (or were not) encoded as part of a schema^39^. Activity here is reduced in people with cognitive impairment^48^ and correlates with behavioural task performance^49^. The IPL more broadly is considered a convergence zone of multiple brain networks that support the actualisation of cognitive operations across both lower-level (i.e. spatial attention) and higher-level (i.e. semantic memory) cognitive functions^50^. Although the left IPL is generally considered more important for language and semantic processing, damage of the left IPL clinically leads to issues contextualising the meaning of objects, events and situations^51^, a key component of the processing required for our task. Furthermore, bilaterally the IPL is a major hub of the multiple association network, the default mode network (DMN), which has been particularly relevant to the action of 5-HT_4_ R agonists in previous studies^23, 52^.

In contrast with our previous prucalopride studies, there were no clear effects of PF-04995274 on behavioural measures of learning and memory^22^. In particular, we did not replicate previous findings of improved recognition of previously seen images in the fMRI memory task or improved recall in the Auditory Verbal Learning Task. There was, however, an interesting effect of PF-04995274 on the Oxford Memory Task, where participants in the active condition were faster to respond, whilst maintaining a similar level of accuracy as the placebo group. This is consistent with the findings from a recent study in our group that investigated the effects of prucalopride in people who had recovered from depression, in which the 5-HT_4_R agonist acted to speed performance across a number of cognitive tasks^53^. Overall, however, PF-04995274 did not have a strong effect on behavioural measures of memory, which suggests that it may have a more subtle pro-cognitive effect than prucalopride that is only evident using sensitive neuroimaging outcome measures. This is the first study assessing the potential pro-cognitive effects of a 5-HT_4_R agonist in a clinical population. An important strength of RESTAND was that our participants with depression were not on any antidepressant treatment, enabling assessment of PF-04995274 without interference from other serotonergic medication. Perhaps partially related to the need for participants to be medication free, the average HAM-D score was in the mild to moderate range at 16, although severity of depression and severity of cognitive difficulties are poorly correlated^54^.

Interestingly, the clinician rated depression scores (HAM-D) were lower in the PF-04995274 group compared with placebo at the end of the study; both in the full set of participants undertaking behavioural testing^25^ and in this cognitive fMRI subset. This is consistent with previous pharmaco-epidemiological work examining the reduced incidence of first diagnoses of depression in the following year after a prescription of the highly-selective 5-HT_4_ agonist, prucalopride, for constipation when compared to alternative anti-constipation agents^55^. However, findings from previous studies as well as other analyses from this study^25^ indicate that 5-HT_4_ R agonism may not influence emotional processing in a typical way as seen with first-line antidepressants^23^, and thus any effect on mood is likely to be occurring via an alternative mechanism. For example, 5-HT_4_R agonists may affect other neural factors associated with depression, such as modulating activity within the hippocampus^22^ and connectivity within the default mode network^23, 52^. However, the effects of 5-HT_4_R agonism on depression scores seen in this sample should be treated cautiously, given this study was not designed or powered to measure clinical efficacy endpoints, and not powered to investigate correlations between brain measures and questionnaire measures.

Whilst this study was not adequately powered to examine sex differences, we includes sex as a regressor in fMRI analyses since overall activity in the hippocampal mask varied according to sex, and we examined the effect of sex in behavioural analyses. The pre-clinical work establishing the optimal dose for receptor occupancy was only conducted in men, and it is possible men and women may show differences in terms of receptor binding and processing of PF-04995274. Additionally, a previous study demonstrated that use of oral contraceptives in females led to lower 5-HT_4_R binding globally, with the largest difference present in the hippocampus^56^. Approximately one third of the females within each group were recorded to be taking systemic contraception (six out of 15 females taking placebo; five out of 17 females taking the novel 5-HT_4_), but these small numbers precluded meaningful sub-analysis.

In conclusion, this study demonstrates that PF-04995274, a new 5-HT_4_R agonist, increases hippocampal function, replicating our previous findings in a clinical population and using a different 5-HT_4_R agonist. This is a critical step in the translation of the considerable preclinical evidence establishing the 5-HT_4_R as a relevant target for the treatment of depression and cognition. Future work is needed to further examine the effects of 5-HT_4_R agonism on clinical efficacy endpoints in order to fully realise the clinical potential of this target.

## Data Availability

All data produced in the present study are available upon reasonable request to the authors. Code is available at OSF.

## Acknowledgements

We would like to thank numerous research assistants for their support with data collection (Lucy Wright, Ingrid Martin, Evie Watson) and data processing (Esther Teo), medics for support with clinical cover (Michael Browning, Riccardo De Giorgi, Tarek Zghoul, Paul Harrison, Sara Costi), Cassandra Gould Van Praag for her support with the scanning protocol and analysis pipelines, numerous staff at the Oxford Health Clinical Research Facility, and the Oxford Health BRC Patients Active in Research Group who provided consultation on study adverts and protocol. We would also like to extend thanks to all of the participants who took part in the study, especially as this involved specifically giving their time and energy during current depressive episodes. We thank Nico Filippini and Masud Husain respectively for the original design of the fMRI memory task and the Oxford Memory Task used in this paper, and Orla MacDonald for her pharmaceutical support. We would like to thank Pfizer for the novel agent used in this study (see Conflict of interest and funding).

## Code availability

Analysis code is available at the Open Science Framework (https://osf.io/v7kgs/?view_only=1cd5bb7f964f445397ba16dc72cfb812).

## Conflict of interest and funding

This study was funded by the Medical Research Council (MRC): MR/S035591/1 and Pfizer under the MRC asset sharing scheme. This study has been delivered through the National Institute for Health and Care Research (NIHR) [Oxford Health] Biomedical Research Centre (BRC) including the Oxford Health cognitive health Clinical Research Facility, and the Oxford Centre for Human Brain Activity (OHBA) part of the Wellcome Centre for Integrative Neuroimaging. The views expressed are those of the author(s) and not necessarily those of the MRC, the NIHR or the Department of Health and Social Care.

ANdeC is funded by an NIHR Clinical Lectureship and also receives funding and support from the NIHR Mental Health Translational Research Collaboration (MH-TRC) Mental Health Mission and the NIHR Oxford Health Biomedical Research Centre. She has previously received funding from the Guarantors of Brain and a Wellcome Trust Clinical Doctoral Research Fellowship (216430/Z/19/Z). ALG, MAGM and WH also are supported by the NIHR Oxford Health Biomedical Research Centre. BG is also supported by the NIHR Mental Health Translational Research Collaboration (MH-TRC) Mental Health Mission. CJH has received consultancy fees from P1vital Ltd., Jannsen Pharmaceuticals, UCB, Compass Pathways, and Lundbeck. She is a co-director of TnC Psychiatry and Neuroscience. SEM has received consultancy fees from Zogenix, Sumitomo Dainippon Pharma, P1vital Ltd. and Johnson & Johnson Pharmaceuticals. CJH and SEM recently held grant income from Zogenix, UCB Pharma and Janssen Pharmaceuticals and ADM. CJH, SEM and PJC recently held grant income from a collaborative research project with Pfizer. ALG has received consultancy fees from Zogenix and Johnson & Johnson Pharmaceuticals. AG and JC have no funding or conflicts of interest to declare.

## Notes

### Competing Interest Statement

This study was funded by the Medical Research Council (MRC): MR/S035591/1 and Pfizer under the MRC asset sharing scheme.
CJH has received consultancy fees from P1vital Ltd., Jannsen Pharmaceuticals, UCB, Compass Pathways, and Lundbeck. She is a co-director of TnC Psychiatry and Neuroscience. SEM has received consultancy fees from Zogenix, Sumitomo Dainippon Pharma, P1vital Ltd. and Johnson & Johnson Pharmaceuticals. CJH and SEM recently held grant income from Zogenix, UCB Pharma and Janssen Pharmaceuticals and ADM. CJH, SEM and PJC recently held grant income from a collaborative research project with Pfizer. Other authors have no conflicts of interest to declare.

### Clinical Trial

NCT03516604

### Author Declarations

The study was approved by the South Central Research Ethics Committee (18/SC/0076) and pre-registered with clinicaltrials.gov (NCT03516604).

