## Supplementary Material for "Effects on hippocampal activity following 5-HT4 receptor agonism in unmedicated patients with depression: the RESTAND study"

### Supplementary Methods

##### Table S1: Exclusion criteria for RESTAND

History of or current DSM-5 bipolar disorder, schizophrenia or eating disorders, or clinically significant risk of suicide

First-degree relative with a diagnosis of Bipolar Disorder type 1

Current usage of psychotropic medication

Failure to respond to antidepressant medication in current episode

Electroconvulsive therapy for the treatment of the current episode of depression

Participants undergoing any form of face-to-face structured psychological treatment during the study

Clinically significant abnormal values for screening blood tests or ECG

History of stimulant abuse (lifetime; e.g. amphetamine, cocaine), or of alcohol abuse within one year or of alcohol dependence within the lifetime

History of, or current medical conditions which may interfere with the safety of the participant or the scientific integrity of the study, including epilepsy/seizures, brain injury, severe hepatic or renal disease, severe gastro-intestinal problems, severe neurological problems

Medical conditions that may alter the hemodynamic parameters underlying the BOLD signal (e.g., inadequately treated hypertension, diabetes mellitus). Any contraindication to MRI scanning (e.g. metal objects in body, pacemakers, significant claustrophobia, pregnancy)

Pregnancy, planning a pregnancy, or breastfeeding

Participants with Body Mass Index (BMI) outside the 18 to 36 kg/m2 range at the Screening Visit

Night-shift working or recent travel involving significant change of time zones, or excessive caffeine consumption

Participation in a psychological or medical study involving the use of medication within the last 3 months

#### *Participants*

90 right-handed participants between 18 and 61 years old were recruited to the full RESTAND study between August 2018 and July 2022. Participants were of either sex, fluent in English, and were screened for contraindications to serotonergic medication. All participants met DSM-5 criteria for major depressive disorder at screening, as determined by a psychiatrist using the Structured Clinical interview for DSM-5 (SCID). Other criteria for inclusion included: being willing and able to give consent; not receiving drug or face-to-face psychological treatment for the last six weeks; and, where relevant, using a highly effective method of contraception from study enrolment until 30 days after study treatment. Participants who had failed to respond to an antidepressant or required ECT in this current episode of depression were excluded, as well as those with diagnoses of bipolar disorder or at high risk of suicide (see Table S1 for a complete list of all exclusion criteria).

The study was approved by the South Central Research Ethics Committee (18/SC/0076) and a protocol including outcomes was pre-registered with clinicaltrials.gov (NCT03516604). Participants gave written informed consent and were paid £200 for their participation. The study was adapted for COVID-19 precautions, but no other changes occurred after the start of recruitment.

#### *Design and randomisation*

##### Overview of design and randomisation

Participants were randomised to one of three groups: PF‐04995274 (15mg), citalopram (20mg) or placebo, in a double-blind, randomised, between-groups design, stratified by gender. Oxford Health Foundation Trust Pharmacy (Clinical Pharmacy Support Unit (CPSU), Kennington) was responsible for drawing up and holding the randomisation list (using an online randomisation tool (Sealed Envelope)). Group allocation was concealed from participants, investigators and assessors using sequential numbered containers. PF‐04995274 and identically-matched placebo tablets were provided by Pfizer. Citalopram and identically-matched placebo capsules were sourced from and encapsulated by Cardiff and Vale University Health Board, St. Mary's Pharmaceutical Unit (SMPU). Participants were instructed to take medication in the morning for up to 9 days, which contained either one active tablet (PF‐04995274 or citalopram) and one placebo, or two sets of placebo tablets.

##### Study visits

The study included four visits to the Warneford Hospital, University of Oxford Department of Psychiatry, with scanning occurring at the Oxford Centre for Human Brain Activity (OHBA) part of the Wellcome Integrative Neuroimaging Centre (WIN): (i) Screening Visit; (ii) First Dose Visit; (iii) Research Visit One (including the fMRI scan) on day 6-9, and (iv) Research Visit Two (including behavioural cognitive tasks and end of study assessments) on day 7-9.

During screening, all participants underwent a medical and psychiatric evaluation including a review of medical history, current and past medication, weight, height and BMI measurement, vital signs assessment, 12-lead ECG, clinical laboratory blood tests for standard screening (liver function tests, urea and electrolytes), urine drug screening, and a pregnancy test where relevant. All participants completed an MRI screening safety form and handedness test to check for MRI eligibility. Following screening, the participant’s GP was informed of their participation in the study.

As the study drug (PF-04995274) was unlicensed, the first study dose was administered under medical supervision within four weeks of screening, with vital signs monitored for three hours afterwards. Participants took the remaining days of medication at home. During the week of drug / placebo administration, participants were advised not to drink alcohol and not to carry out activities requiring full alertness if they were aware of any impairment. A researcher also contacted them by phone on day two and day four to check that there were no concerns, and participants received a daily text message reminding them to take the study medication. From day six, all participants were invited for an MRI scan. Spare medication was given to all participants to enable completion of study visits up to and including day nine to allow for scheduling difficulties or illness. The dosing of PF‐04995274 used here (15 mg for at least 6 days) is within the previously studied dosing and duration of administration carried out by Pfizer where preliminary studies found it to be safe and generally well tolerated (Pfizer 2011b), with a half-life (t_1/2_) of around 30 hours and greater than 80% occupancy of brain 5-HT_4_Rs four hours after the first dose in healthy volunteers (Pfizer 2011a).

##### Sample Size

A sample size calculation was based on data acquired in (25) comparing citalopram to placebo, aiming for 0.9 power and a 0.05 false positive rate. This suggested a group sample size of 19 per group for off-line behavioural analyses. As 5-HT_4_R agonism is less well studied, it was aimed to recruit at least 25 individuals per group to ensure adequate power for fMRI analyses based on active intervention (PF‐04995274 OR citalopram) versus placebo group comparisons. Based on the hippocampal region of interest results from (24), this sample size should be sufficient to reliably detect an effect size of the same magnitude (G*Power, previous effect size (f) = 0.37, with an α=0.05, to give 90% power (1-β)). The citalopram group was included as a secondary comparison versus placebo for emotional cognition analyses (explored elsewhere). We therefore include citalopram versus placebo results for reference only in this supplement.

#### *Questionnaire measures*

Participants completed the following self-report questionnaires at Screening to obtain measures of trait anhedonia, anxiety, and personality: Snaith–Hamilton Pleasure Scale (SHAPS) (26), Spielberger State-Trait Anxiety Inventory, Trait Version (STAI-T) (27) and Eysenck Personality Questionnaire (28). Affect and state anxiety were also measured pre- and post-imaging (Research Visit 1): affect using the Positive and Negative Affect Scale (PANAS) (29) and the visual analogue scale (VAS) (30), and for anxiety the Spielberger State-Trait Anxiety Inventory, State Version (STAI-S) (27). Side effects were measured at Screening and pre- and post-imaging using a scale where participants rated the extent to which they were experiencing each of the most reported side effects of prucalopride (31). Participants also assessed on an observer-rated Hamilton Rating Scale for Depression (HAM-D) by trained researchers and completed a self-report measure of depressive symptoms (Beck Depression Inventory-II (BDI))(32) at Screening and Research Visit Two. At the end of Research Visit Two, participants also guessed their drug allocation with a multiple-choice question.

#### *Memory encoding fMRI task*

This was conducted exactly as detailed in (24) during Research Visit 1. Outside and before the scan, participants were shown eight randomly-selected “familiar” pictures (four animals and four landscapes). Participants were asked to identify these as animal / non-animal. During the scanner, these “familiar” images were shown in blocks of eight images alternating with blocks of “novel” images (six familiar and six novel blocks) interleaved with rest blocks (a cross on the screen). In each task block, eight images were presented in a pseudorandom order: either eight “familiar” images (the images presented prescan) or eight “novel” images (previously unseen images). Familiar images were presented in a different order in each block. Novel images were taken from a pool of 48 images (24 animals and 24 landscapes) and each was only presented once during the task. During the scan, participants again had to identify these images as animal / non-animal using a two-button response and were informed there would be a memory test afterwards. After the scan, these eight “familiar” and 48 “novel” images were shown alongside 27 “distractor” images, and participants were asked if they had seen the image in the scanner or not. 83 images were presented on a PC screen for 4000 ms each (interstimulus interval 1000 ms): the eight “familiar” images (seen prescan and during scan), the 48 “novel” images (seen during scan only), and 27 “distractors” (images never seen before: 13 animals and 14 landscapes) were displayed in pseudorandom order. The task was programmed in Presentation (Neurobehavioral Systems; [https://www.neurobs.com](https://www.neurobs.com/)). This task was designed to stimulate implicit visual memory recognition in the scanner for images that participants had seen before (familiar images), compared to images that were new to participants in the scanner (novel images) where implicit encoding should occur.

#### *Auditory verbal learning task (AVLT)*

The AVLT was conducted as part of Research Visit 2. Participants were read a list of 15 concrete nouns (List A) and asked to immediately verbally recall as many items as they could. This was repeated five times before a list of unrelated words was presented (List B) and participants were again asked to recall them. Participants were then asked to recall List A immediately (short-delay) and after a delay of ∼15 min (long-delay). Number of words correct, repetitions (correct words recalled more than once in the same acquisition trial) and intrusions (incorrect words not present in the list) were measured. Participants then completed a recognition task where they were required to indicate which of a list of words (15 List A words, 35 distractors) had previously been presented. Number of hits and false alarms were measured.

#### *Demographic and behavioural data analysis*

Behavioural data and questionnaires were processed and analysed using SPSS (version 29, IBM). Graphs were produced using GraphPad Prism (version 9) and R (version 4.3.3). A repeated-measures analysis of variance (ANOVA) was used to analyse group differences in self-report measures and behavioural performance in the AVLT and fMRI experimental task. Levene’s test (t-tests) and the Greenhouse–Geisser procedure (ANOVAs) were used where appropriate. A p-value less than 0.05 was used to denote statistical significance. Partial eta squared is reported as a measure of effect size. As the RESTAND population were a clinical population from across adulthood, age, sex, and HAM-D score were considered as potential confounds for behavioural task analysis.

#### *MRI data acquisition*

fMRI acquisition

Encoding memory was assessed from a single run of 60 T2-weighted echoplanar imaging (EPI) slices covering the whole brain [repetition time (TR) 800 ms, echo time (TE) 30 ms, flip angle 52°, field of view 216 mm, slice thickness 2.4mm, voxel dimension 2.4mm isotropic, acquisition time 6min 48s]. Images were distortion corrected by an acquired fieldmap (echos at 4.92 and 7.38 ms, TR=590ms, flip angle = 46°).

Structural MRI acquisition

Additional high-resolution T1-weighted structural scans were acquired using a gradient echo sequence (TR 1900ms, TE 3.97ms, flip angle 8°, field of view 192mm, voxel dimension 1 mm isotropic, acquisition time 5min 31s) to allow later registration of the fMRI data into standard space.

ASL acquisition

Each participant also had a resting state pCASL perfusion-weighted scan with a 2D gradient spin echo readout and a PICORE Q2T labelling scheme. ASL data were collected as tag-control pairs with a TI of 1.8 seconds and a bolus duration of 0.7 seconds. ASL imaging parameters were: repetition time: 4100ms; minimum echo time: 14.0ms; FOV read: 220mm; FOV phase: 100%; voxel size: 3.4x3.4x4.5mm; 24 slices with 4.5mm thickness; echo spacing: 0.56mm; EPI factor: 64; post-labelling delays at: 250ms, 500ms, 750ms, 1000ms, 1250ms, and 1500ms; number of dynamics/repeats: 97 (1 volume was control); acquisition time = 6min 39s; fat saturation = on. A calibration image was acquired without labelling (TR = 6000ms).

Labelling plane was set with a time of flight neck scan (TR = 21ms, TE = 3.43ms, flip angle = 30°, field of view = 200mm, voxel dimension = 0.3 x 0.3 x 1.3 mm, acquisition time = 42s). Images were distortion corrected by an acquired fieldmap (echos at 4.92 and 7.38ms, TR=482ms, flip angle = 46°).

#### *MRI analysis*

fMRI task analysis: Imaging data were analysed with FSL ([www.fmrib.ox.ac.uk/fsl](http://www.fmrib.ox.ac.uk/fsl)).

fMRI data were pre-processed and analysed using FEAT (FMRI Expert Analysis Tool), version 6.0.4, part of FSL (FMRIB’s Software Library; www.fmrib.ox.ac.uk/fsl). DICOM (Digital Imaging and Communications in Medicine) files were downloaded from the server, checked for completeness, excessive movement and visual anomalies, and converted to a BIDS (Brain Imaging Data Structure)-standardised format nifti files using heudiconv (heudiconv 0.5.4 (<https://github.com/nipy/heudiconv)>) before pre-processing. The structural anatomical scans were brain extracted using the Brain Extraction Tool (BET)^1^.

Pre-processing of functional data involved: motion correction using FMRIB’s Linear Image Registration Tool (FLIRT^2^); deletion of non-brain tissue using BET ^1^; spatial smoothing with a Gaussian kernel of 5 mm full-width-half-maximum; grand-mean intensity normalisation of the entire 4D dataset by a single multiplicative factor; high pass temporal filtering (Gaussian-weighted least-squares straight line fitting, with sigma of 90s) and B0 unwarping using fieldmap phase and magnitude images for distortion correction. No slice timing correction was applied (temporal derivates were included in the model to account for differences in slice timing). In addition, registration to high-resolution image and to a standard template [Montreal Neurological Institute (MNI)] was implemented using FNIRT nonlinear registration ^3^.

In the first-level analysis, individual activation maps were computed using the general linear model with local autocorrelation correction. Two explanatory variables were modelled: “novel” and “familiar” images. Temporal derivatives were included in the model. Variables were modelled by convolving each block with a haemodynamic response function, using a variant of a gamma function (standard deviation 3s, mean lag 6s). No included participant demonstrated significant movement: absolute displacements were less than 1 voxel and relative displacements less than ½ voxel. At the whole-brain level, familiar images were contrasted with novel, resulting in the following model: 1) novel vs. baseline; 2) familiar vs. baseline; 3) novel > familiar; 4) novel < familiar.

In the second-level analysis, whole-brain individual data were combined at a group level (participants on placebo vs. 5-HT_4_) using a mixed-effects analysis, and cerebral blood flow and grey matter maps as covariates of no interest. Groups were contrasted with each other, resulting in the following comparisons: 1) placebo > 5-HT_4_; 2) 5-HT_4_> placebo; 3) placebo mean; 4) 5-HT_4_mean; 5) mean of all participants. Brain activations showing significant group differences were identified at the whole-brain level using cluster-based thresholding (Z>3.1, family-wise error (FWE) p<0.05 corrected). Significant interactions from whole-brain analyses were further explored by extracting percentage BOLD signal change for each type of contrast. As the hippocampus was a particular focus, it was pre-specified as a region of interest (ROI). A functional ROI mask was created for the left and right hippocampus by multiplying mean activation for each contrast of interest (on whole-brain data already corrected for multiple comparisons (FWE) and Z>3.1 as described above) for all participants by the Harvard-Oxford subcortical atlas anatomical mask at a 50% threshold. Percentage BOLD signal change for each contrast in each hemisphere was extracted in order to identify the profile of drug effect. All activations are reported using MNI co-ordinates.

FSLVBM, a voxel-based morphometry style analysis ^4^, was carried out to investigate potential grey matter differences between the two study-groups, underlying and potentially influencing group-related BOLD differences. Brain-extracted images (automatically created for each individual using FSLanat) were tissue-type segmented. Grey matter partial volume images were aligned to standard space using FLIRT and FNIRT registration tools. The resulting images were averaged, modulated and smoothed with an isotropic Gaussian kernel of 2mm to create a study-specific-template. A voxel-wise GLM was then applied using permutation non-parametric testing (5000 permutations).

Regional and global blood flow was calculated for each individual. Distortion and motion corrected resting perfusion maps in units of mL/100g/min were calculated using Oxford_ASL (part of the Bayesian Inference for Arterial Spin Labelling (BASIL) tool, https://fsl.fmrib.ox.ac.uk/fsl/fslwiki/BASIL; ^6, 7^ for each participant, which performs label-control subtraction, inference of voxelwise perfusion, and voxelwise calibration to obtain absolute perfusion maps, and controls for partial volume effects at the single subject level. FSL’s Anatomical Processing Script (FSL_Anat, https://fsl.fmrib.ox.ac.uk/fsl/fslwiki/fsl_anat) was used to pre-process each participant’s high resolution T1 structural image (includes bias-field correction, brain extraction and registration to standard space via FMRIB’s Linear Image Registration Tool (FLIRT) and FMRIB’s Non-linear Image Registration Tool (FNIRT). The processed perfusion images were non-linearly aligned with standard space via an initial linear transformation T1 structural space (using FLIRT), followed by application of the non-linear warp from fsl_anat. A Gaussian smoothing kernel of 2.12mm was applied to all the normalised images (to match functional data). Data were interrogated using voxel-wise generalized linear model (GLM) permutation nonparametric testing (5,000 permutations) with randomise (FSL’s tool for nonparametric permutation inference on neuroimaging data), correcting for multiple comparisons across space (cluster-based thresholding using TFCE and a family-wise error (FWE)-corrected cluster significance threshold of p<0.05 applied to the suprathreshold clusters). This results in spatial maps characterising the between-subject/group differences.

Hippocampal perfusion between groups was compared using fslmeants: parameter estimates of perfusion were extracted from resting perfusion maps (previously computed using Oxford_ASL in units of ml/100g/min) using anatomical Harvard-Oxford masks of the left and right hippocampus at a 50% threshold.

Similar to (24), for fMRI analyses, we pre-specified to report results with grey matter and perfusion correction, and considered other covariates such as sex as relevant.

### Supplementary Results

##### Figure S1: CONSORT diagram

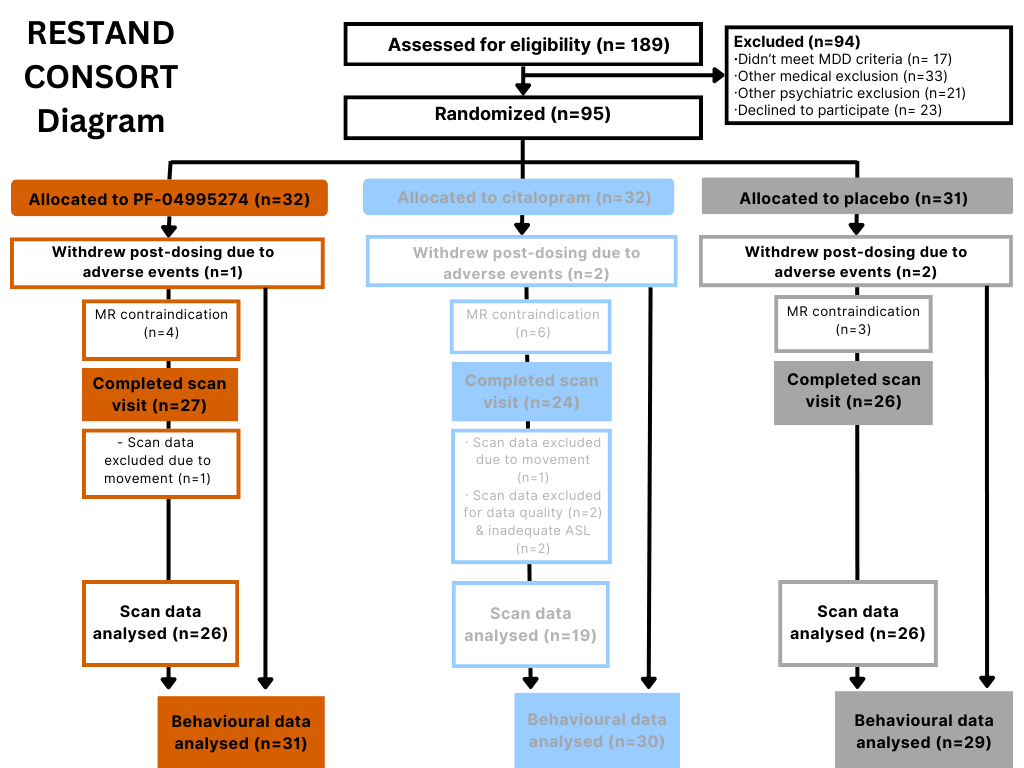

This CONSORT diagram is similar to Gillespie et al and specific to this paper in terms of data exclusions. The citalopram arm was an active comparison for the emotional processing outcomes reported in Gillespie et al, but main outcomes from this paper comparing citalopram and placebo are detailed here in the Supplementary Material.

##### Table S2: Medical history of included participants

|  | **Placebo** | **5HT_4_** |
| --- | --- | --- |
| ***Eye disorders*** | 1 (managed cataract) | 0 |
| ***Musculoskeletal symptoms*** | 1 | 2 |
| ***Previous mild concussion*** | 1 | 1 |
| ***Autoimmune diseases*** | 1 (hypothyroidism (corrected)) |  |
| ***Mild asthma*** | 0 | 2 |
| ***History of kidney stones*** | 0 | 2 |
| ***Hayfever*** | 0 | 2 |
| ***Previous headaches / migraines*** | 0 | 2 |
| ***Dyspepsia*** | 0 | 3 |
| ***Gynaecological disorders*** | 1 (endometriosis) |  |

##### Table S3: Current medication history of included participants

|  | **Placebo** | **5-HT_4_** |
| --- | --- | --- |
| ***Analgesics (paracetamol / ibuprofen / aspirin / codeine)*** | 5 | 4 |
| ***Proton pump inhibitors (omeprazole / lansoprazole)*** | 2 | 1 |
| ***Levothyroxine*** | 1 | 0 |
| ***Antihistamines*** | 0 | 2 |
| ***Statins*** | 0 | 1 |
| ***Inhalers*** | 0 | 2 |

##### Table S4: Results of randomisation guesses at the end of the study

|  | **Placebo**  **N=25*^[1]^** | **PF-04995274**  **N=26** |
| --- | --- | --- |
| ***Incorrect guess*** | 7 | 9 |
| ***Correct guess*** | 11 | 3 |
| ***Uncertain*** | 7 | 14 |

##### Table S5: State anxiety and affect questionnaire results during the scan visit (placebo: 5-HT_4_)

|  | **Placebo mean (SD)**  **N=26** | **PF-04995274 mean (SD)**  **N=24*^[2]^** | **P value (2dp)** |
| --- | --- | --- | --- |
| **Spielberger State Anxiety Inventory (STAI-S)**  ***Pre-Testing***  ***Post-Testing*** | 44.8 (9.54)  44.2 (10.1) | 43.3 (11.0)  40.4 (10.6) | 0.35 |
| **Positive and Negative Affect Scale –**  **Positive (PANAS-P)**  ***Pre-Testing***  ***Post-Testing*** | 22.3 (6.44)  22.5 (8.48) | 22.1 (8.56)  23.1 (7.74) | 0.48 |
| **Positive and Negative Affect Scale –**  **Negative (PANAS-N)**  ***Pre-Testing***  ***Post-Testing*** | 17.2 (6.23)  14.9 (5.44) | 15.1 (5.38)  14.1 (4.73) | Included in ANOVA above |
| **Visual Analogue Scale (VAS) –**  **Happy**  ***Pre-Testing***  ***Post-Testing*** | 42.3 (17.3)  45.5 (22.6) | 44.7 (22.5)  50.5 (20.6) | 0.37 |
| **Visual Analogue Scale (VAS) –**  **Sad**  ***Pre-Testing***  ***Post-Testing*** | 38.9 (19.9)  31.2 (28.9) | 32.3 (28.8)  26.8 (24.6) | Included in ANOVA above |
| **Visual Analogue Scale (VAS) –**  **Hostile**  ***Pre-Testing***  ***Post-Testing*** | 13.2 (14.3)  13.2 (18.4) | 13.1 (18.0)  12.2 (16.9) | Included in ANOVA above |
| **Visual Analogue Scale (VAS) –**  **Alert**  ***Pre-Testing***  ***Post-Testing*** | 46.0 (21.0)  39.5 (24.3) | 40.0 (24.0)  35.0 (22.1) | Included in ANOVA above |
| **Visual Analogue Scale (VAS) –**  **Anxious**  ***Pre-Testing***  ***Post-Testing*** | 45.4 (20.6)  34.0 (29.8) | 35.1 (29.7)  26.4 (27.8) | Included in ANOVA above |
| **Visual Analogue Scale (VAS) –**  **Calm**  ***Pre-Testing***  ***Post-Testing*** | 49.6 (22.4)  49.2 (22.1) | 48.5 (22.0)  56.9 (20.2) | Included in ANOVA above |

#### Supplementary behavioural results of the fMRI memory encoding task

##### Table S6: ANOVA results for distinguishing each category of image (novel/familiar/distractor) in behavioural fMRI task, controlling for age, baseline or change in HAM-D, sex, or when including the participant excluded for movement from fMRI analyses

|  | **F statistic** | **P value** |
| --- | --- | --- |
| ***+ Age*** | F(1,44)=0.126 | 0.725 |
| ***+ Baseline HAM-D scores*** | F(1,44)=0.231 | 0.633 |
| ***+ Age + Baseline HAM-D scores*** | F(1,43)=0.282 | 0.598 |
| ***+ HAM-D score change**** | F(1,43)=0.034 | 0.854 |
| ***+ Sex*** | F(1,44)=0.151 | 0.699 |
| ***+ participant excluded for movement*** | *F*(1,46)=0.016 | 0.901 |

* = final HAM-D scores missing for 1 participant in 5HT4 group

Results were essentially unchanged when we also ran a sensitivity analysis excluding a statistical outlier from the analysis (also in the PF-04995274 group) (i.e.) for distinguishing each category of image (novel/familiar/distractor): F(1,44)=0.001, *p*=0.973 (Placebo: PF-04995274 = 25:21).

*Main analyses for the behavioural part of the memory encoding task repeated for citalopram (n=19) versus placebo (n=26):*

In the post-scan recall test, there was no difference between groups in accuracy to identify images seen before (novel + familiar) versus distractors [*F*(1,41)=1.09, *p*=0.30, ηρ2=0.03; overall mean for placebo (M=77.2% SEM=1.82), citalopram (M=79.5%, SEM 2.30), or when distinguishing each category of image (novel/familiar/distractor) [F(1,41)=0.11, *p*=0.74, ηρ2<0.01]. There was no interaction between image type (novel/familiar/distractor) and group [F(1.4,57.6)=2.15, *p*=0.14, ηρ2=0.05)]. Data missing for 2 participants (1 from placebo, 1 from citalopram = 18:25).

#### Supplementary results for grey matter analyses

There were group-related differences in grey matter between the placebo and 5-HT_4_ groups (FSLVBM randomise: placebo > 5-HT_4_, p=0.03; 5-HT_4_ > placebo, p=0.99) involving the left lateral occipital cortex [placebo > 5-HT_4_, peak voxel location X=-28,-80,26, cluster size = 43 voxels]. As pre-specified, whole-brain and hippocampal region of interest analyses include grey matter correction, but were similar with and without.

##### Figure S2: Group-related grey matter differences between placebo and 5-HT_4_ groups (FSLVBM)

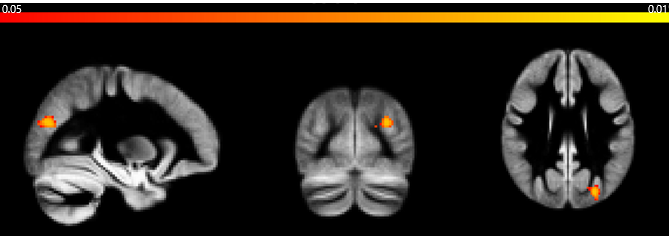
*Sagittal, coronal, and axial images for FSLVBM at MNI location 45,54,45. Images thresholded at p<0.05. Red to yellow colours identify increases in grey matter differences (see colour bar included in image)*

#### Supplementary results in terms of MRI perfusion

There was no difference between placebo and PF-04995274 groups in regional blood flow (randomise: placebo > 5-HT_4_, *p*=0.75; 5-HT_4_ > placebo, *p*=0.21) or global blood flow (Oxford_ASL: *p*=0.40 (grey matter); *p*=0.24 (white matter)). Left and right hippocampal perfusion did not differ between groups (*p*=0.14 (LHC smoothed); *p*=0.16 (RHC smoothed) (see Table S6)). As pre-specified, whole-brain and hippocampal region of interest analyses include cerebral perfusion correction, but were similar with and without.

##### Table S7: Hippocampal perfusion (smoothed and unsmoothed) across the right and left hemisphere (ml/100g/min)

|  |  | **Left Hippocampus** | **Right Hippocampus** |
| --- | --- | --- | --- |
| **Smoothed** | ***Placebo*** | 50.4 | 50.8 |
|  | ***5-HT_4_*** | 53.7 | 53.8 |
| **Unsmoothed** | ***Placebo*** | 51.7 | 52.5 |
|  | ***5-HT_4_*** | 55.0 | 55.6 |

#### Supplementary results for memory encoding task fMRI

##### Figure S3: Regions with increased BOLD fMRI signal intensity for main effect of task (novel > familiar)

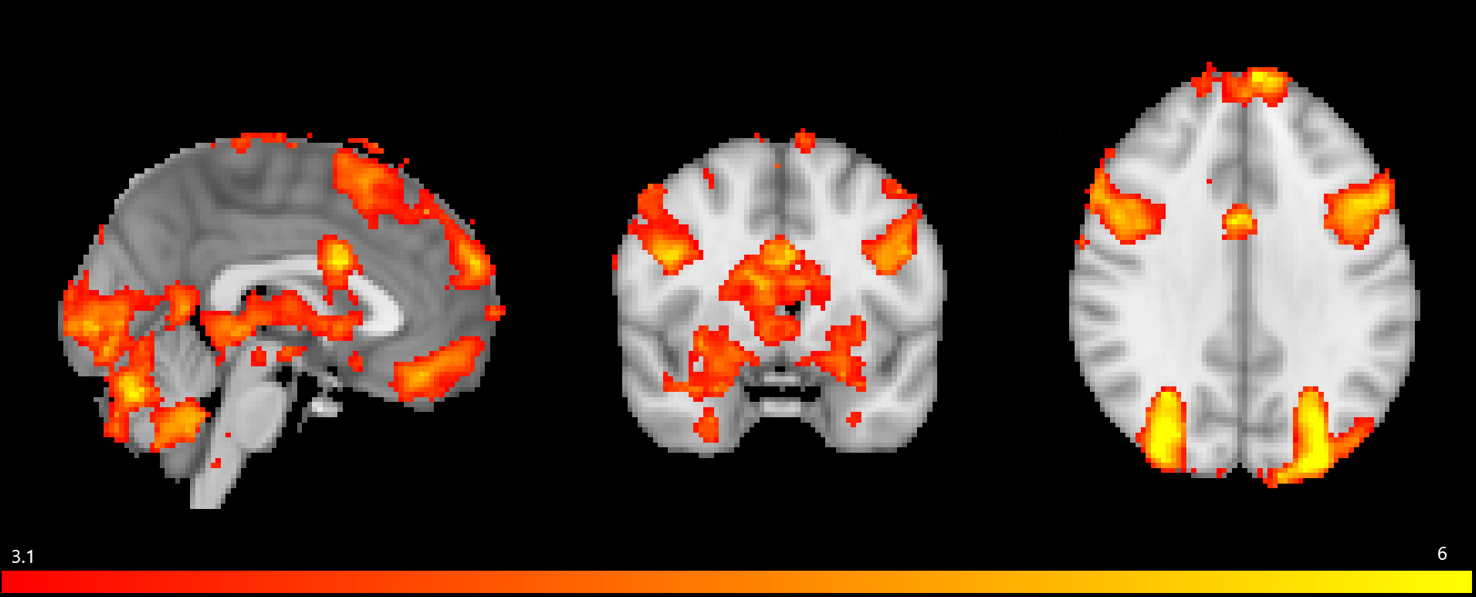

*Sagittal, coronal, and axial images for the novel vs. familiar contrast at MNI location 45,65,52. Images thresholded at z > 3.1. Red to yellow colours identify increases in brain activation (see colour bar included in image)*

##### ***Effect of treatment – region of interest analysis***

Condition individually was also run in a similar ANOVA as a secondary analysis: LHC familiar vs RHC familiar: F(1,49)=1.81, p=0.18, np2=0.04; LHC novel vs RHC novel: F(1,49)=1.06, p=0.31, np2=0.02.

(LHC = left hippocampus; RHC = right hippocampus)

*Main analyses for the fMRI memory encoding task repeated for citalopram (n=19) versus placebo (n=26):*

In the hippocampal region of interest mask, there was no significant main effect of group [F(1,41)=0.06, *p*=0.80, ηρ2<0.01] or significant condition*hemisphere*group interaction [F(1,41)=0.26, *p*=0.61, ηρ2=0.01].

##### ***Effect of treatment – whole brain analysis***

Results for when ASL+GM+sex were included as covariates (for direct comparison with ROI analyses): [5-HT_4_>placebo, Z=3.9, p<0.014, peak voxel location: X=-36, Y=-82, Z=36, cluster size = 131 voxels].

Placebo and 5-HT_4_ groups differed in grey matter in this region (placebo > 5-HT_4_). Of note, cluster is still present here when whole-brain analyses were run without correction for grey matter.

Whole brain results were essentially identical to main analyses when controlled for baseline HAM-D and age.

#### Supplementary results for AVLT

##### Table S8: List A immediate recall score results by group for AVLT

| **Block** | **Placebo (N=29)** | **PF-04495274 (N=29)** |
| --- | --- | --- |
| ***A – Time 1*** | 7.28 (1.87) | 7.49 (1.82) |
| ***A – Time 2*** | 10.4 (2.54) | 10.5 (2.5) |
| ***A – Time 3*** | 12.3 (2.06) | 12.3 (2.36) |
| ***A – Time 4*** | 12.7 (2.13) | 12.9 (1.89) |
| ***A – Time 4*** | 13.3 (2.00) | 13.1 (2.07) |

##### Table S9: ANOVA results for AVLT List A Short and Long Delay, List B, and Repetitions and Intrusions

|  | **F statistic** | **P value** |
| --- | --- | --- |
| ***List B*** | F(1,56)=0.47 | 0.49 |
| ***Short Delay List A*** | F(1,56)=0.00 | 0.96 |
| ***Long Delay List A*** | F(1,55)=0.13 | 0.72 |
| ***Intrusions*** | F(1,56)=0.00 | 1.00 |
| ***Repetitions*** | F(1,56)=0.20 | 0.66 |

There was a lower mean recall for males compared to females in the PF-04995274 group, but not in the placebo group for List B / List A Short Delay / List A Long Delay. Males compared to females in the PF-04995274 group also had a higher rate of repetitions and intrusions (see Table S11).

##### Table S10: Mean (SD) recall by group when sex was considered

|  | **Placebo (N=26)** | | **PF-04495274 (N=26)** | |
| --- | --- | --- | --- | --- |
|  | **Males** | **Females** | **Males** | **Females** |
| ***List B*** | 6.83 (1.95) | 6.47 (2.24) | 5.30 (1.49) | 87.90 (1.82) |
| ***Short Delay List A*** | 11.0 (3.57) | 12.2 (2.46) | 8.90 (2.33) | 13.2 (1.72) |
| ***Long Delay List A*** | 11.3 (3.14) | 12.2 (2.80) | 8.8 (3.01) | 13.1 (2.13) |
| ***Intrusions*** | 1.33 (1.50) | 1.35 (1.58) | 2.70 (2.45) | 0.63 (0.96) |
| ***Repetitions*** | 4.92 (3.37) | 4.65 (4.12) | 7.90 (7.03) | 3.90 (2.85) |

There were also no differences in results when we included the participant excluded from fMRI analyses for movement, or when we accounted for age or HAM-D scores (see Table S12).

##### Table S11: ANOVA results for AVLT List A recall controlling for age, baseline or change in HAM-D

|  | **F statistic** | **P value** |
| --- | --- | --- |
| ***+ Age*** | F(1,54)=0.48 | 0.50 |
| ***+ Baseline HAM-D scores*** | F(1,54)=0.56 | 0.46 |
| ***+ Age + Baseline HAM-D scores*** | F(1,50)=0.03 | 0.86 |
| ***+ HAM-D score change**** | F(1,53)=0.17 | 0.68 |
| ***+ Sex*** | F(1,54) = 0.48 | 0.49 |

* = final HAM-D scores missing for 1 participant in 5HT4 group

*Main analyses for AVLT repeated for citalopram (n=30) versus placebo (n=29):*

There was a significant effect of block on word recall [F(7,399) = 178, p < 0.001, np2=0.76], indicating that in both groups participants’ recall of words from List A improved across the five acquisition blocks. However, there was no significant interaction between the block and group [F(7, 399) = 0.48, p = 0.85, np2<0.01] or main effect of group [F(1,57) = 0.08, p = 0.77, np2<0.01].

#### Supplementary results for OMT

##### Table S12: Mean (SD) results for OMT for 1 fractal and 3 fractals

(A) Fractals 1:

|  | **5HT4** | | **Placebo** | |
| --- | --- | --- | --- | --- |
|  | **Mean** | **SD** | **Mean** | **SD** |
| ***Accuracy*** | 87.41 | 10.79 | 90.54 | 8.96 |
| ***Identification Time*** | 2331.97 | 738.80 | 2343.48 | 609.03 |
| ***Localisation Time*** | 4263.62 | 1190.64 | 4713.24 | 1190.99 |

(B) Fractals 3:

|  | **5HT4** | | **Placebo** | |
| --- | --- | --- | --- | --- |
|  | **Mean** | **SD** | **Mean** | **SD** |
| ***Accuracy*** | 97.58 | 4.06 | 97.86 | 4.99 |
| ***Identification Time*** | 1176.02 | 353.92 | 1238.49 | 386.93 |
| ***Localisation Time*** | 2845.01 | 725.12 | 3073.48 | 578.45 |

##### Table S13: Estimated marginal means and pairwise comparisons for localisation time model with covariates

(A) WITH AGE AS COVARIATE

Estimated marginal means

| **Fractals** | **Group** | **emmean** | **SE** | **df** | **Lower CL** | **Upper CL** |
| --- | --- | --- | --- | --- | --- | --- |
| ***fractal1*** | Placebo | 4782 | 183 | 82.7 | 4418 | 5145 |
| ***fractals3*** | Placebo | 3142 | 183 | 82.7 | 2779 | 3505 |
| ***fractal1*** | 5-HT_4_ | 4217 | 174 | 81.9 | 3869 | 4564 |
| ***fractals3*** | 5-HT_4_ | 2798 | 174 | 81.9 | 2451 | 3145 |

Degrees-of-freedom method: kenward-roger

Confidence level used: 0.95

Pairwise comparisons

| **Contrast** | **Estimate** | **SE** | **df** | **T ratio** | **P value** |
| --- | --- | --- | --- | --- | --- |
| ***fractal1_LocalRT Placebo - fractals3_LocalRT Placebo*** | 1640 | 174 | 60.1 | 9.398 | <.0001 |
| ***fractal1_LocalRT Placebo - fractal1_LocalRT 5-HT_4_*** | 565 | 255 | 81.8 | 2.220 | 0.1264 |
| ***fractal1_LocalRT Placebo - fractals3_LocalRT 5-HT_4_*** | 1984 | 255 | 81.8 | 7.792 | <.0001 |
| ***fractals3_LocalRT Placebo - fractal1_LocalRT 5-HT_4_*** | -1074 | 255 | 81.8 | -4.220 | 0.0004 |
| ***fractals3_LocalRT Placebo - fractals3_LocalRT 5-HT_4_*** | 344 | 255 | 81.8 | 1.352 | 0.5332 |
| ***fractal1_LocalRT 5-HT_4_ - fractals3_LocalRT 5-HT_4_*** | 1419 | 166 | 60.1 | 8.555 | <.0001 |

Degrees-of-freedom method: kenward-roger

P value adjustment: tukey method for comparing a family of 4 estimates

(B) WITH SEX AS COVARIATE

Estimated marginal means

| **Fractals** | **Group** | **emmean** | **SE** | **df** | **Lower CL** | **Upper CL** |
| --- | --- | --- | --- | --- | --- | --- |
| ***fractal1*** | Placebo | 4695 | 187 | 81.4 | 4323 | 5067 |
| ***fractals3*** | Placebo | 3055 | 187 | 81.4 | 2683 | 3427 |
| ***fractal1*** | 5-HT_4_ | 4223 | 180 | 80.3 | 3866 | 4581 |
| ***fractals3*** | 5-HT_4_ | 2805 | 180 | 80.3 | 2448 | 3162 |

Results are averaged over the levels of: Sex

Degrees-of-freedom method: kenward-roger

Confidence level used: 0.95

Pairwise comparisons

| **Contrast** | **Estimate** | **SE** | **df** | **T ratio** | **P value** |
| --- | --- | --- | --- | --- | --- |
| ***fractal1_LocalRT Placebo - fractals3_LocalRT Placebo*** | 1640 | 174 | 60.1 | 9.407 | <.0001 |
| ***fractal1_LocalRT Placebo - fractal1_LocalRT 5-HT_4_*** | 471 | 256 | 81.5 | 1.843 | 0.2610 |
| ***fractal1_LocalRT Placebo - fractals3_LocalRT 5-HT_4_*** | 1890 | 256 | 81.5 | 7.389 | <.0001 |
| ***fractals3_LocalRT Placebo - fractal1_LocalRT 5-HT_4_*** | -1168 | 256 | 81.5 | -4.567 | 0.0001 |
| ***fractals3_LocalRT Placebo - fractals3_LocalRT 5-HT_4_*** | 250 | 256 | 81.5 | 0.978 | 0.7621 |
| ***fractal1_LocalRT 5-HT_4_ - fractals3_LocalRT 5-HT_4_*** | 1419 | 166 | 60.1 | 8.563 | <.0001 |

Results are averaged over the levels of: Sex

Degrees-of-freedom method: kenward-roger

P value adjustment: tukey method for comparing a family of 4 estimates

(C) WITH HAM-D (RV2) AS COVARIATE

Estimated marginal means

| **Fractals** | **Group** | **emmean** | **SE** | **df** | **Lower CL** | **Upper CL** |
| --- | --- | --- | --- | --- | --- | --- |
| ***fractal1*** | Placebo | 4754 | 187 | 80.3 | 4381 | 5126 |
| ***fractals3*** | Placebo | 3114 | 187 | 80.3 | 2741 | 3486 |
| ***fractal1*** | 5HT4 | 4182 | 184 | 77.6 | 3815 | 4549 |
| ***fractals3*** | 5HT4 | 2769 | 184 | 77.6 | 2402 | 3136 |

Degrees-of-freedom method: kenward-roger

Confidence level used: 0.95

Pairwise comparisons

| **Contrast** | **Estimate** | **SE** | **df** | **T ratio** | **P value** |
| --- | --- | --- | --- | --- | --- |
| ***fractal1_LocalRT Placebo - fractals3_LocalRT Placebo*** | 1640 | 176 | 59.1 | 9.325 | <.0001 |
| ***fractal1_LocalRT Placebo - fractal1_LocalRT 5-HT_4_*** | 572 | 268 | 77.7 | 2.137 | 0.1506 |
| ***fractal1_LocalRT Placebo - fractals3_LocalRT 5-HT_4_*** | 1985 | 268 | 77.7 | 7.420 | <.0001 |
| ***fractals3_LocalRT Placebo - fractal1_LocalRT 5-HT_4_*** | -1068 | 268 | 77.7 | -3.993 | 0.0008 |
| ***fractals3_LocalRT Placebo - fractals3_LocalRT 5-HT_4_*** | 345 | 268 | 77.7 | 1.290 | 0.5718 |
| ***fractal1_LocalRT 5-HT_4_ - fractals3_LocalRT 5-HT_4_*** | 1413 | 170 | 59.1 | 8.319 | <.0001 |

Degrees-of-freedom method: kenward-roger

P value adjustment: tukey method for comparing a family of 4 estimates

##### Figure S4: OMT scores for PF-04495274 (5HT4) and placebo groups post-intervention for OMT

(A) Localisation time by age

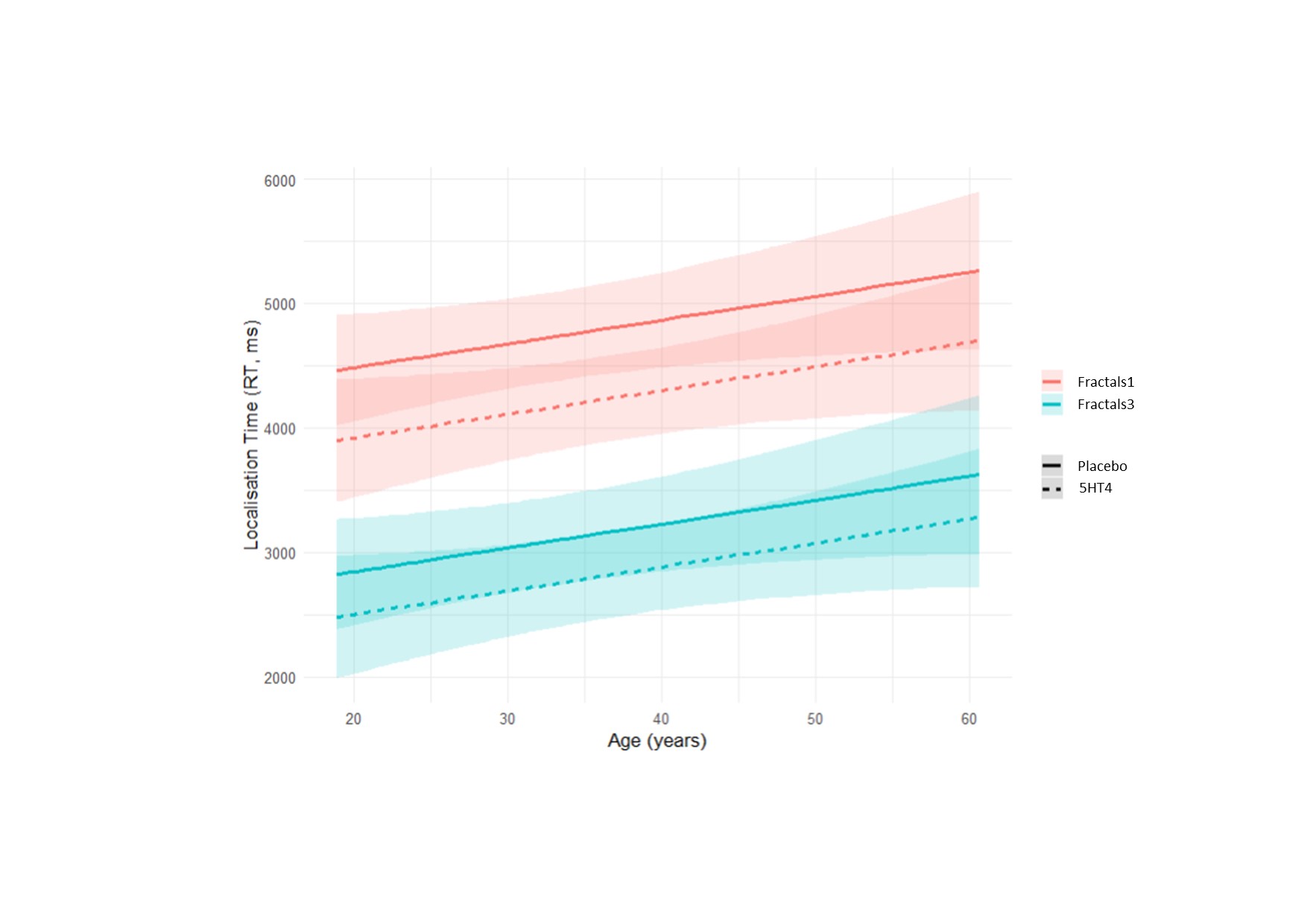

(B) Localisation time by HAM-D score

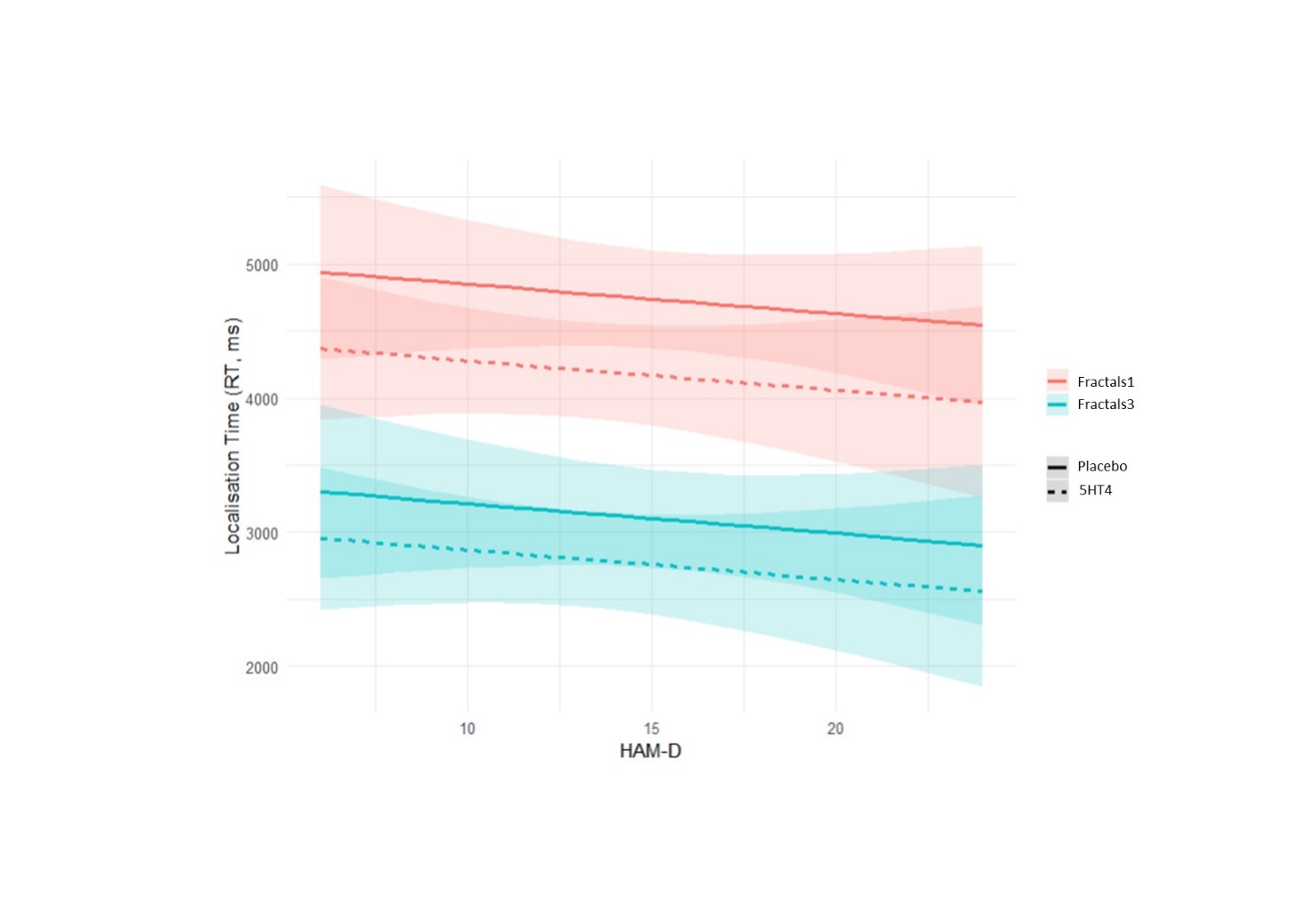

(C) Localisation time by sex

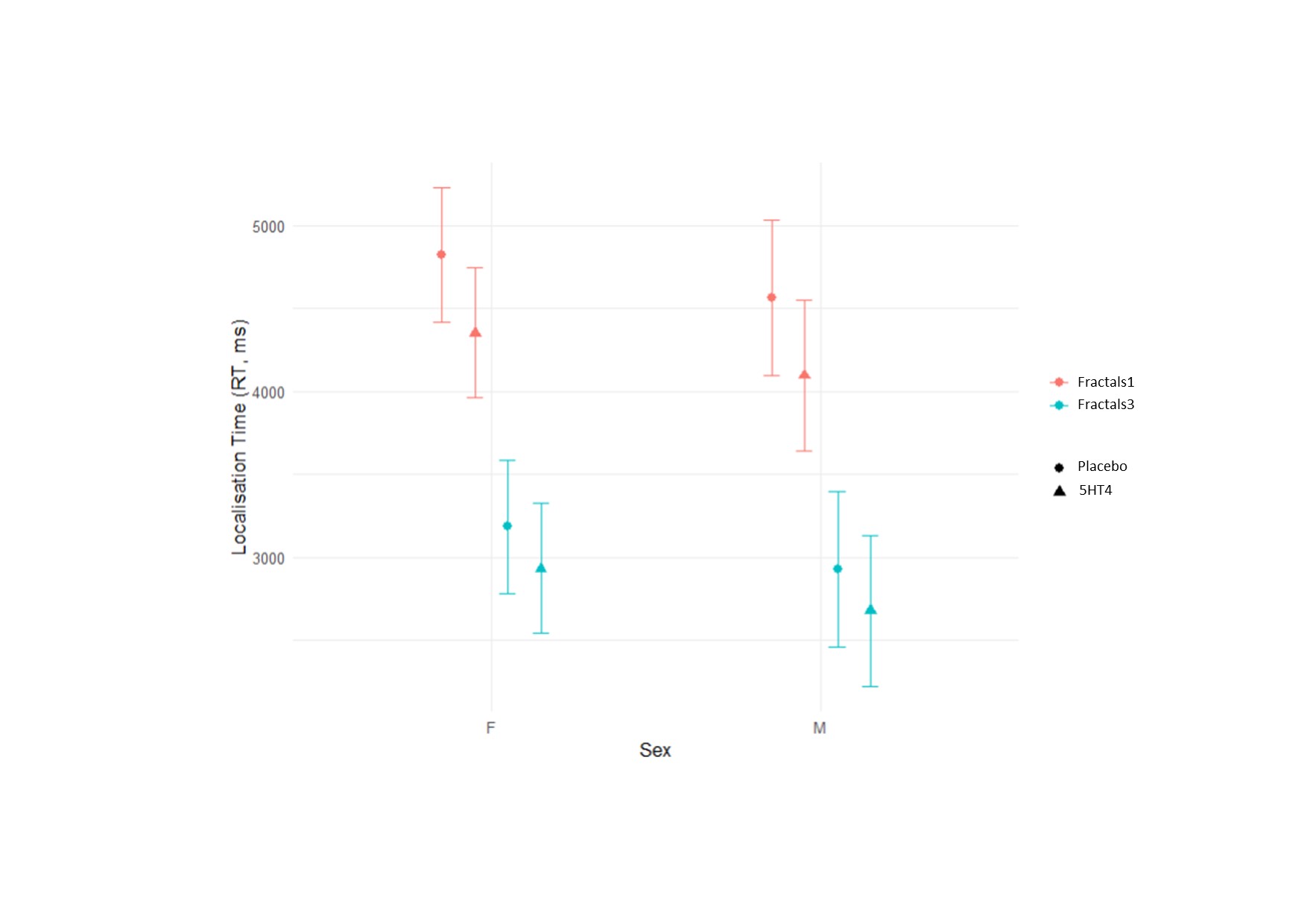

M= males; F = females; RT = reaction time, ms = milliseconds.

*Main analyses for OMT repeated for citalopram (n=29) versus placebo (n=28):*

There was no difference between groups in terms of accuracy [main effect of group (F(1,53.0)=0.65, p = 0.42, np2 = 0.01); fractals***group** (F(1,55.3)=1.90, p = 0.17, np2 = 0.03)], or identification time [main effect of group (F(1,52.6)=0.23, p = 0.64, np2 < 0.01); fractals***group** (F(1,55.0)=0.01, p = 0.90, np2<0.01)].

Contrasting to the 5HT4/placebo comparison, there was no difference on localisation time [main effect of group (F(1,53.7)=0.9, p = 0.34, np2 = 0.02); fractals***group** (F(1,55.8)=0.55, p = 0.46, np2<0.01)].

1. 1 participant missing in placebo group
2. 2 participants missing from 5-HT_4_ group
